# Pandemic-response adenoviral vector and RNA vaccine manufacturing

**DOI:** 10.1101/2021.08.20.21262370

**Authors:** Zoltán Kis, Kyungjae Tak, Dauda Ibrahim, Maria M Papathanasiou, Benoît Chachuat, Nilay Shah, Cleo Kontoravdi

## Abstract

Rapid global COVID-19 pandemic response by mass vaccination is currently limited by the rate of vaccine manufacturing. This study presents a techno-economic feasibility assessment and comparison of three vaccine production platform technologies deployed during the COVID-19 pandemic: (1) adenovirus-vectored (AVV) vaccines, (2) messenger RNA (mRNA) vaccines, and (3) the newer self-amplifying RNA (saRNA) vaccines. Besides assessing the baseline performance of the production process, the impact of key design and operational uncertainties on the productivity and cost performance of these vaccine platforms were also evaluated using variance-based global sensitivity analysis. Cost and resource requirement projections were also computed for manufacturing multi-billion vaccine doses for covering the current global demand shortage and for providing annual booster immunizations. This model-based assessment provides key insights to policymakers and vaccine manufacturers for risk analysis, asset utilisation, directions for future technology improvements and future epidemic/pandemic preparedness, given the disease-agnostic nature of these vaccine production platforms.

## 1. Introduction

The COVID-19 pandemic, caused by the SARS-CoV-2 virus, created an unprecedented demand for rapid, large-scale vaccine deployment that the world is struggling to meet. This urgency and scale of immunization against a new disease poses enormous challenges on the entire vaccine deployment pipeline [1–4]. This pipeline has the following main parts: 1) pre-clinical development and testing, 2) clinical development and testing, 3) production process development, scale-up and technology transfer for the manufacturing of the vaccine active ingredient (drug substance, DS), 4) sourcing of raw materials and consumables for manufacturing both the drug substance and the final packaged vaccine product filled into glass vials or other containers (fill-to-finish processes, f2f), 5) drug substance production under current Good Manufacturing Practices (cGMP), 6) fill-to-finish processes under cGMP, 7) vaccine distribution and 8) vaccine administration to the population [1–4].

COVID-19 vaccination programmes around the world are currently limited by the number of vaccine doses that can be manufactured [5]. To pre-emptively address this challenge and reduce deployment timelines, setting up vaccine manufacturing for pandemic-response production started “at risk” before the safety and efficacy of the vaccines was confirmed in clinical trials [4,6]. This has the following challenges: (a) uncertainty in the DS amount per dose and number of doses per person, which are determined during clinical trials, (b) production processes have to be developed, optimised and scaled up [1]. Additionally, new vaccine designs may be required to tackle new virus variants. Finally, manufacturing needs to be low-cost to enable mass immunization, including in low- and middle-income countries [1,7].

Herein, we review the manufacturing processes for adenovirus-vectored (AVV), messenger RNA (mRNA), and the newer self-amplifying RNA (saRNA) vaccines. These vaccines contain genetic instructions, in the form of DNA for the AVV vaccine and RNA in case of the mRNA and saRNA vaccines, based on which the cells of the human body produce the vaccine antigen, such as the spike protein of the SARS-CoV-2 virus [8–13]. Because only the genetic instruction and not the antigen is produced, the vaccine production processes serve as platform technologies. A platform technology implies that once validated and established at production scale, the same production processes can produce a wide range of different vaccines and vaccine candidates against both known and currently unknown, future, pathogens. The AVV and mRNA vaccine platforms have matured in terms of technology development and have high technology readiness levels. On the other hand, the saRNA vaccine platform is currently in clinical development and has a low technology readiness level.

We then analyse key uncertainties andtheir impact on COVID-19 vaccine production, as well as quantifying the production process scales, timescale and manufacturing resources required for producing 1 billion COVID-19 vaccines per year. These estimates can serve as a basis for calculating the requirements to produce vaccines for the global demand. Following drug substance production, we further evaluate three fill-to-finish technologies with respect to their pandemic-response manufacturing performance: conventional fill-to-finish in 5-dose or 10-dose vials, blow-fill-seal in single-dose vials, and the new 200-dose bag Intact™ Modular Filler [1,14–16].

All the models included in this study are representative of industrially-relevant cGMP COVID-19 vaccine manufacturing processes [17,18]. The technologies used for COVID-19 vaccine production and their productivity in terms of number of vaccine doses produced per unit time and unit scale of the process varies tremendously among the three technologies. In order to compare them on the same basis, the following key performance indicators (KPIs) have been used: (i) annual production amounts expressed in doses per year, and (ii) and cost per dose expressed in USD per dose. The values of these KPIs and the degree by which these are impacted by the uncertainties was also assessed using global sensitivity analysis [19–21]. The rate at which batches can be produced and the number of doses produced per batch was also evaluated, as completing production batches in a short space of time would yield a steadier vaccine supply which can be advantageous in an emergency response situation compared to a more fragmented supply characteristic to production batches that require a longer time to complete. This study can inform policy makers and vaccine manufacturers on how to improve manufacturing and asset utilisation against COVID-19 and its variants, but also against future outbreaks due to the disease-agnostic nature of these vaccine production platforms [1,22].

## 2. Results and Discussion

### 2.1. Comparative technological assessment of COVID-19 vaccine production platforms

The AVV, mRNA and saRNA DS production processes (primary manufacturing) are described in the SI document. Fill-finish processes are described in **Table S1**. Likely baseline scenarios are presented in **Table S2**, and one-factor-at-a-time uncertainty analysis in **Figure S1** shows the impact of scale, titre and DS amount per dose on annual production amounts and production costs.

In reality, these input parameters may vary simultaneously. Thus, we have carried out variance-based stochastic global sensitivity analysis [23–25] to evaluate how input uncertainty propagates to outputs and aportion it individual inputs and their interactions [23–25], as illustrated in **Figure 1**.

**Figure 1.**
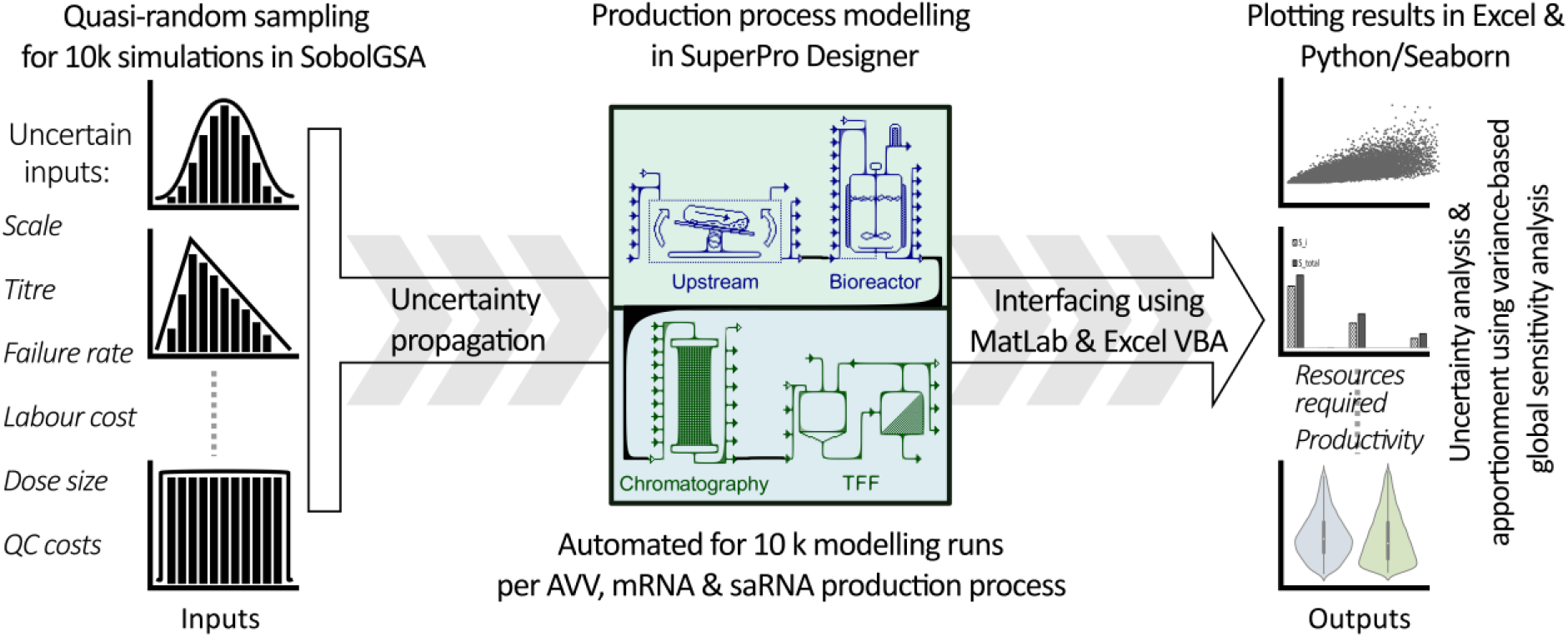
Graphical illustration of the computational framework for uncertainty quantification. The aim of this approach is to evaluate process performance under uncertainty and variability resulting from both the design and operation of the new vaccine production platform technologies. The uncertainty is propagated from the inputs via the model to the outputs. In addition, the sensitivity of the model output key performance indicators (KPIs) is attributed to the individual inputs to determine the degree to which individual inputs impact the output KPIs. Model inputs include the scale of the production process, batch failure rate, titre/yield in the production bioreactor, cost of labour, drug substance amount per dose and cost of quality control. Model outputs include capital investment cost requirements, operating costs, number of batches produced per year, amount of drug substance produced per batch, amount of drug substance produced per year, number of doses produced per batch, number of doses produced per year and the production cost per dose.

The uncertain input variables and corresponding ranges for the three platforms are listed below in **Table 1** and with additional explanation on the factors influencing these ranges in **Table S4** of the SI document. The probability of the uncertainty distribution was set to triangular when the probable value was considered to have a high probability, and set to uniform when the input values in the range were considered to have equal or similar probability of occurring. For each platform technology 10,000 production process models were simulated using quasi-randomly generated combinations of model input variables within the ranges shown below in **Table 1**. Sensitivity and uncertainty analyses were not performed for the fill-to-finish process as these are well-established technologies (relevant values are shown in Table S1).

**Table 1.** Input variables for sensitivity analysis for mRNA, saRNA and AVV vaccine drug substance production modelling.

| Parameter name and unit | AVV |  | mRNA & saRNA (RNA) |  | Uncertainty Distribution* | Ref. |
| --- | --- | --- | --- | --- | --- | --- |
|  | Range | Central Value** | Range | Probable Value |  |  |
| Process scale [L for RNA and AVV] | 1000 – 20000 | 2000 | 0.5 – 50 | 30 mRNA<br>5 saRNA | Triangular | [26–29] |
| Process failure rate [%] | 0 – 10 | 5 | 0 – 10 | 5 | Uniform | [30] |
| Production titres [g L<sup>-1</sup> for RNA; viruses L<sup>-1</sup> for AVV] ** | $1 \times 10^{14} - 7 \times 10^{14}$ | $2.5 \times 10^{14}$ | 3 – 7 | 5 | Triangular | [26,27] |
| 5' cap analogue cost [USD g<sup>-1</sup>] | N.A. | N.A. | 2500 – 10000 | 3000 | Triangular | [31] |
| Basic labour rate [USD hour<sup>-1</sup>] | 5 – 30 | 20 | 5 – 30 | 20 | Triangular | [30] |
| Drug substance amount per dose [µg dose<sup>-1</sup> for RNA; viruses dose<sup>-1</sup> for AVV] | $2.2 \times 10^{10} - 6.5 \times 10^{10}$ | $5 \times 10^{10}$ | 0.1 – 10 saRNA<br>5 – 150 mRNA | 1 saRNA<br>100 mRNA | Triangular | [32–44] |
| Cost of Lab/QC/QA [% of total labour costs] | 20 – 60 | 40 | 15 – 60 | 40 | Uniform | [30] |

\* Uncertainty distribution was considered either triangular, with highest probability for the central value, or uniform, with equal probability for all the values in the range.
\*\* These are the values which were used in the baseline scenarios. For parameters which have a triangular distribution, the central value also corresponds to the mode. For uniform distributions the central value has the same frequency/abundance as the other values in the range.
\*\*\* The changes in recovery rate or losses in the downstream purification were modelled by changing the product titre in the bioreactor, as the ranges of changes in product titre also account for variations in amount of drug substance produced per batch, which could be due to changes in downstream purification losses.

Each of the scatter plots in **Figure 2** show the impact of one input parameter onto one output parameter. The global sensitivity analysis results for AVV show that the annual production amounts are mostly influenced by scale, followed by titre and then AVV amount per dose as shown by the height of the bars in **Figure 2G**. The AVV cost per dose is mostly impacted by uncertainty in scale, titre, and AVV amount per dose, however the contribution of these inputs is of comparable magnitude as shown in **Figure 2H** and **Figure 2D-F**. The annual production amounts for both mRNA and saRNA depend mostly on the RNA amount per dose followed by the production scale, as shown in **Figure 2O&I-J** and **Figure 2W&Q-S** for mRNA and saRNA, respectively. The cost per dose for both mRNA and saRNA is predominantly influenced by the RNA amount per dose and to a relatively lesser extent by the production titres and the price of the CleanCap 5’ cap analogue, as shown in **Figure 2P&L-N** and **Figure 2X&T-V** for mRNA and saRNA, respectively. The differences between the magnitude of the S_t_ and S_i_ indicate that all process models have predominantly separable or additive characteristics with regards to the impact of the inputs on the outputs. The input parameters which have the highest impact on the outputs align with the width of the ranges of the input parameters. The width of these ranges is associated with the specifics of the platform technologies and the corresponding technology readiness levels.

**Figure 2.**
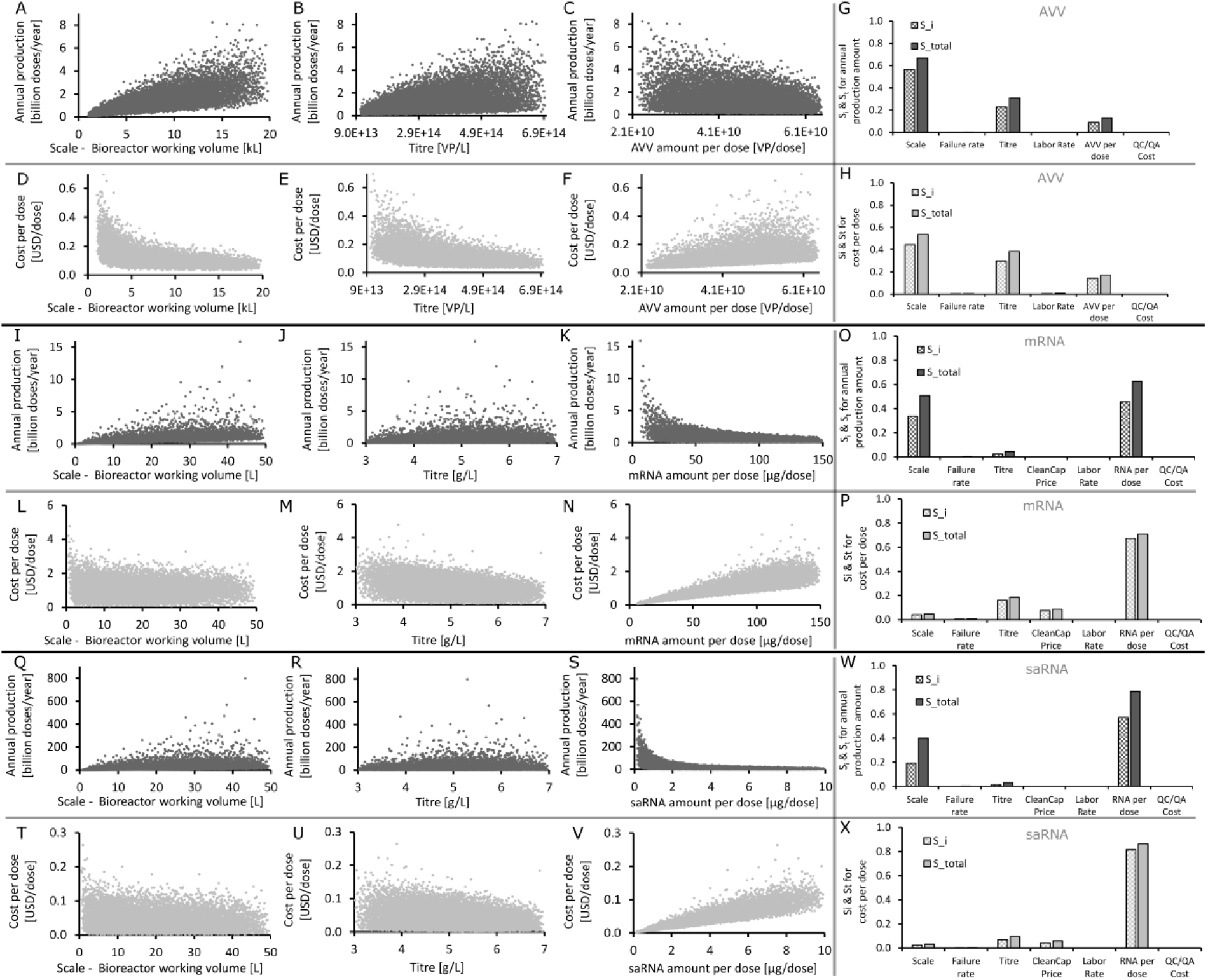
The impact of co-variation of the input parameters on the annual production amounts and cost per dose of AVV, mRNA and saRNA drug substance production captured by stochastic global sensitivity analysis using quasi-random sampling based on Sobol sequences [23–25]. A-H (top two rows), I-P (middle two rows), Q-X (bottom two row) shows AVV, mRNA and saRNA vaccine drug substance production performance, respectively. **A-F**. Scatter plots showing the random co-variation of AVV drug substance annual production amounts in function of production scale, titre and AVV amount per dose. **G-H**. The main-effect (aka. 1st-order effect) Sobol indexes (S_i_) and total-order effect Sobol indexes (S_t_) plotted in function of the seven input parameters shown on the x-axis for AVV drug substance production global sensitivity analysis. **I-N**. Scatter plots showing the random co-variation of mRNA drug substance annual production amounts in function of production scale, titre and mRNA amount per dose. **O-P**. The S_i_ and S_t_ plotted in function of the seven input parameters shown on the x-axis for mRNA drug substance production global sensitivity analysis. **Q-V**. Scatter plots showing the random co-variation of saRNA drug substance annual production amounts in function of production scale, titre and saRNA amount per dose. **W-X**. S_i_ and S_t_ plotted in function of the seven input parameters shown on the x-axis for saRNA drug substance production global sensitivity analysis. Dots clustered around a narrower region on the Y-axis indicate that the respective input parameter explains most of the variance of the output KPI. On the contrary, dots spread out over a wider region on the Y-axis indicate that the respective input parameter explains little or no variance of the output KPI. Large S_i_ and S_t_ values account for a strong impact of respective inputs shown on the X-axis on the output KPIs on the Y-axis, and low S_i_ and S_t_ values indicate a weaker dependence of the KPIs on the respective inputs.

The productivity of the three platform technologies was compared using variance-based global sensitivity analysis presented above and based on the model input parameter ranges described in **Table 1**. For this comparison, only DS production was modelled and it was assumed that production takes place in one facility and one production line per facility for each of the three vaccine platform technologies. In addition, production processes are assumed to be fully developed, validated and implemented at production scale. The required times (excluding quality control testing) and productivity together with their uncertainty distributions for producing 1 billion doses of DS are shown in **Figure 3**. For this, inside the violin plots, box-and-whisker plots also show the minimum (0th percentile or Q0) and maximum (100th percentile or Q4) values using the extremities of the whiskers and the box plots show the interquartile ranges delimited by the 25th percentile (first quartile or Q1) and the 75th percentile (third quartile or Q3). The median is shown by a white dot inside the box plot, and the outliers are outside the whiskers, thus beyond the minimum and maximum values. The width of violin plots represents the probability distributions.

**Figure 3.**
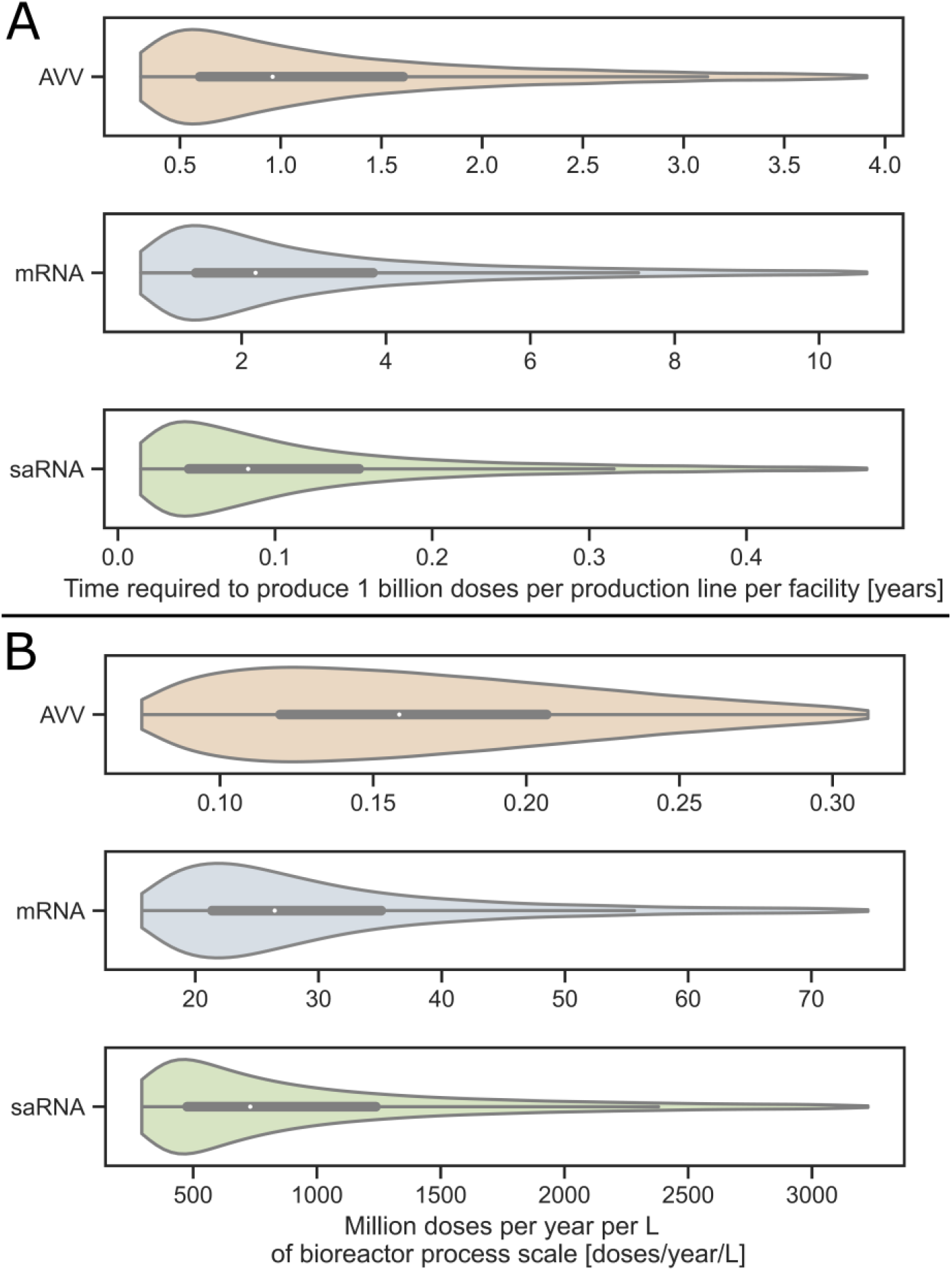
Speed and productivity of the AVV, mRNA and saRNA vaccine production platform technologies. Time requirements for producing drug substance for 1 billion COVID-19 vaccine doses using the AVV, mRNA and saRNA production platforms. **A**. Violin plots showing the computed time requirements for producing 1 billion AVV, mRNA and saRNA vaccine drug substance doses. The time is shown on the x-axis and the production technologies together are listed on the y-axis. **B**. Violin plots showing the number of vaccine doses produced per year per unit of process scale. The unit of process scale is expressed per L of bioreactor working volume. Box and whisker plots are shown in the centre of all violin plots. The box and whisker plots show the minimum and maximum values, except outliers, with the ends of the whiskers; the 25^th^ and 75^th^ percentiles with the top and bottom of the boxes; and the median is shown by the white dot in the box. These global sensitivity analysis results were obtained based on the modelling inputs from **Table 1**. The bottom 5% and top 5% of all values were excluded from all violin plots in order to obtain a better visualization of the region of interest around the box plot. The equivalent violin plots showing all the data (including the top 5% and bottom 5%) are presented in **Figure S3** of the SI document.

As shown in **Figure 3A**, the mRNA platform is likely to require the longest time to produce a unit of 1 billion vaccine doses. More specifically, an mRNA facility with a single production line would require a 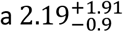 years to produce 1 billion vaccine DS doses. The 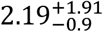 years representation indicates a median of 2.19, the +1.91 superscript is the difference between the 75^th^ percentile and the 50^th^ percentile (which is the mean) and the -0.9 subscript is the difference between the 25^th^ percentile and the 50^th^ percentile. The saRNA platform is more likely to be faster, being capable of producing 1 billion DS doses in 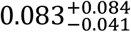 years in a facility with a single production line. However, depending on the uncertainty realisation the saRNA platform may be surpassed by the AVV platform which can produce 1 billion doses in 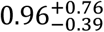 years in a facility with a single production line.

The AVV platform relies on cell-based production which introduces more biological variability which in turn might lead to higher failure rates compared to the mRNA and saRNA platforms, which might further reduce the productivity of the AVV platform. The productivity of the three vaccine platforms, expressed in million doses produced per year per unit scale of the production process (represented by the bioreactor working volume), is shown in **Figure 3B**. Therein, the violin plots show the productivity ranges on the horizontal axis and the vertical width of the violin plots shows the probability density distribution. The mRNA vaccine production process is at 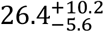 million doses per L of bioreactor working volume per year. As above, the median is shown by the base value, 26.4, the superscript shows the top quartile (75^th^ percentile minus median) and the subscript shows the bottom quartile (25^th^ percentile minus median) values. The productivity of the mRNA platform is two orders of magnitude higher than that of the AVV platform which is at 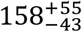 thousands. The saRNA platform at 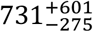 million doses per L of bioreactor working volume per year is one to two orders of magnitude more productive than the mRNA platform and four orders of magnitude more productive than the AVV platform. The productivity difference between the RNA and AVV platforms is due to the highly concentrated, cell-free enzymatic reaction mix based production of the mRNA and saRNA vaccines. The productivity difference between mRNA and saRNA vaccines is due to the substantially lower amount of RNA per dose of the saRNA vaccines.

At the beginning of the production campaign, the start of the fill-to-finish is delayed by the time required to produce and quality test the first DS batch. During pandemic-response manufacturing, the DS can be produced and stockpiled in parallel to carrying out the clinical trials [45], thus the start of fill-to-finish will be even more delayed relative to the start of DS production. The DS production bottlenecks depend on the specific vaccine platform technology. In case of AVV-based vaccines, the bottleneck is caused by the time needed to culture mammalian cells to reach the required amounts in the production bioreactor. In case of mRNA and saRNA vaccine DS production, the bottleneck is in the microfluidics LNP formulation unit operation. The LNP formulation bottleneck can be removed by increasing the size (scaling up) or the number of parallel equipment (scaling out) for the formulation unit operation. However, mRNA vaccine production is most effectively enhanced by reducing the mRNA amount per vaccine dose. The relationship between annual production amount and amount per dose is multiplicative inverse, as shown in **Figure S1H** and **2K**.

Besides bottlenecks in the actual production processes, additional waiting times can be expected for the completion of certain QC tests especially in case of new platform technologies such as the RNA platform. A potential solution for this would be the use of a Quality by Design (QbD) framework to streamline quality assurance by building quality assurance into the design and operation of the production process, which is currently limited by suitable process analytical technology (PAT) [46].

Additionally, the following three different fill-to-finish technologies (secondary manufacturing) processes were also assessed: conventional filling into 10-dose or 5-dose vials, blow-fill-seal into single-dose vials, and the new 200-dose bag Intact Modular Filler system [1,14–16]. For details see SI document and **Table S1**. The overall production bottleneck, when considering both DS production and fill-to-finish, depends on the combination of the specific technologies. For example, in case of filling AVV vaccines in 10-dose vials, the bottleneck will be in the DS production when one AVV DS production line with baseline characteristics (cf. **Table S2**) at the common 2000 L bioreactor working volume scale is coupled to a single 10-dose vial filling line which fills at 400 doses per minute at 60% overall equipment effectiveness (OEE). However, depending on the uncertainty realization (e.g. when larger DS production scales are also considered), the DS production rate might surpass the fill-to-finish rate, as shown by the global sensitivity analysis results presented in **Figure 3A** compared to the values shown in **Table S1**. The baseline mRNA vaccine production rate (cf. **Table S2**) is also slower then filling into 5-dose vials. If an mRNA vaccine production line is coupled with a 10-dose vial (Moderna vaccine) filling line which fills at 400 vials per minute, the overall production bottleneck would again be at the DS production stage. On the other hand, in case of saRNA DS vaccine production coupled with fill-to-finish into 5-dose vials at the 400 vials/minute rate, the overall production bottleneck is at the fill-to-finish stage.

### 2.2. Production process scales and resources required to produce multi-billion doses of Covid-19 vaccine

Considering the above-presented uncertainties, violin plots have been generated to estimate the resource and production capacity requirements for producing a unit of 1 billion doses of vaccine drug substance per year, cf. **Figure 4**. These CapEx, OpEx, production scales and number of batch values as well as their uncertainty distributions were obtained from the variance-based global sensitivity analysis presented in section 2.1 using linear scaling and the model input parameter ranges described in **Table 1**. Linear scaling was used because these production processes and raw material supplies were modelled at large scale, and this would scale approximately linearly when scaling out for meeting the global pandemic demand. The computed CapEx and OpEx requirements for producing 1 billion doses of vaccine drug substance and drug product per year are shown in **Figure 4A** and **Figure 4B**, respectively. For this, it was assumed that AVV vaccines are filled into 10-dose vials, and that both mRNA and saRNA vaccines are filled into 5-dose vials, based on the Oxford/AstraZeneca AVV and the Pfizer/BioNTech mRNA vaccine, respectively. The production scales required to produce a unit of 1 billion doses of AVV, mRNA and saRNA vaccine drug substance per year are shown in **Figure 4C**. The number of AVV, mRNA and saRNA production batches required to produce 1 billion doses of vaccine drug substance per year are shown in **Figure 4D**.

**Figure 4.**
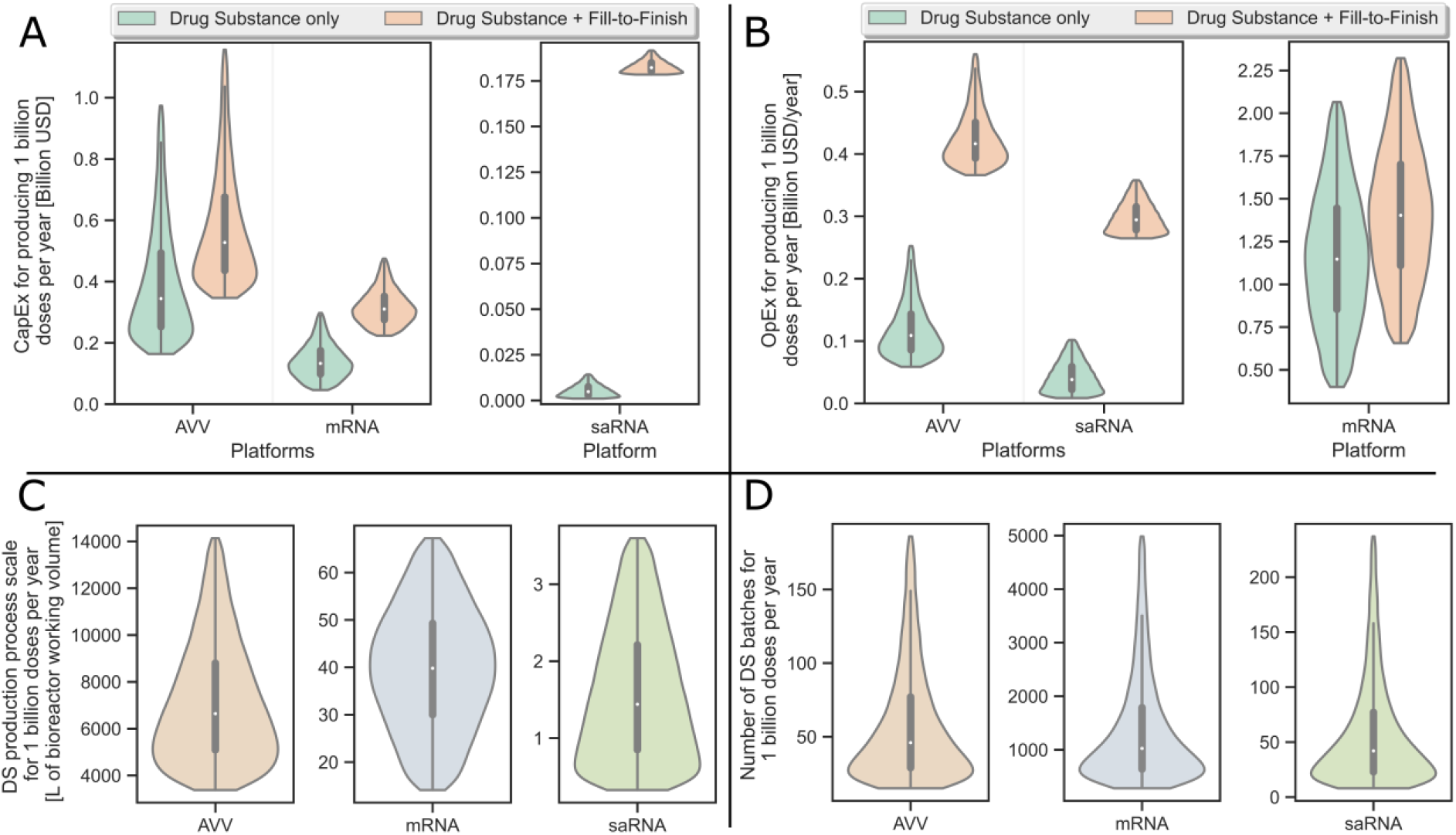
Violin plots showing the global sensitivity analysis of the resource requirements for producing 1 billion COVID-19 vaccine doses per year using the AVV, mRNA and saRNA production platforms combined with conventional liquid fill-to-finish. The inputs and their ranges used for this global sensitivity analysis are shown in Table 1. In the centre of the violin plots, box and whisker plots are shown with the median values indicated by the white dots; the 25th and 75th percentiles with the top and bottom of the boxes; and minimum and maximum values, except outliers, with the ends of the whiskers. **A**. Operating costs (OpEx) required to produce 1 billion doses per year of AVV, mRNA and saRNA vaccine drug substance (DS) and drug product. It was assumed that AVV vaccine is filled into 10-dose vials, whereas the mRNA and saRNA vaccine is filled into 5-dose vials. **B**. Capital costs (CapEx) required to produce 1 billion doses per year of the vaccine drug substance and drug product using the three platform technologies. AVV vaccine fill-to-finish was modelled based on 10-dose vials, whereas the mRNA and saRNA vaccine fill-to-finish was modelled based on 5-dose vials. **C**. Production process scales required to produce 1 billion doses of DS per year using the AVV, mRNA and saRNA vaccine production platforms. The scale of the production process is represented by the working volume in the bioreactor and the entire process is scaled based on the mass balances proportionally to the bioreactor working volume. **D**. Number of batches required to produce 1 billion doses of DS per year using the AVV, mRNA and saRNA vaccine production platforms. The bottom 5% and top 5% of all values were excluded from all violin plots in order to obtain a better visualization of the region of interest around the box plot. The equivalent violin plots showing all the data (including the top 5% and bottom 5%) are presented in Figure S4 of the SI document, which also shows the CapEx and OpEx for drug product manufacturing by fill-to-finish.

Out of the three vaccine drug substance production platform technologies, the mammalian cell-based AVV platform is predicted to have the highest CapEx with a median of ≈340 million USD, with a top quartile (75^th^ percentile of ≈410 million USD minus the median) of +70 million USD and with a lower quartile (25^th^ percentile ≈280 million USD minus the median) of -60 million USD, represented as 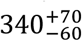 million USD. The AVV platform also requires the highest production scale of 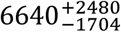 L bioreactor working volume, and second highest OpEx with a 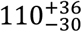 million USD per year to produce a nit of 1 billion vaccine DS doses per year.

Since the AVV platform is commonly implemented at relatively large scales, e.g. at the 2000 L bioreactor working volume scale, it requires a low number of batches to produce 1 billion doses per year, with 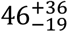 batches per year. The mRNA platform requires the highest number of batches with 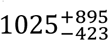 batches, highest OpEx of 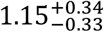 billion USD per year, second highest CapEx of 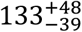 million USD and second highest production scale of 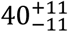 L bioreactor working volume, to produce 1 billion vaccine DS doses. This is due to the high amount of RNA assumed per vaccine dose, with the peak of the triangular distribution representing the Moderna COVID-19 mRNA vaccine, which contains 100 µg of mRNA per dose. However, the mRNA amount per dose can be substantially lower for other mRNA COVID-19 vaccines, for example 30 µg of mRNA per dose for the BioNTech vaccine. The annual mRNA DS production is inversely proportional to the amount per dose (**Figures S1H** & **2K)**. Therefore, the values for the mRNA vaccine production KPIs are expected to improve in case of vaccines with lower mRNA amounts per dose.

The saRNA canditate platform requires the lowest CapEx at 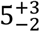 million USD and lowest OpEx of 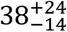 million USD per year to produce 1 billion vaccine doses per year. In addition, the saRNA platform would require the lowest production scales of 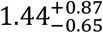 L bioreactor working volume, and depending on the uncertanty realization, possibly the lowest number of batches of 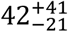 batches to produce 1 billion vaccine doses per year. The OpEx of the mRNA and saRNA vaccines is driven by the high material costs, due to the novelty and limited supply of some of the specialized raw materials required to manufacture mRNA and saRNA vaccines. These materials include the 5’ capping reagents (e.g. 5’ capping analogues such as CleanCap and 5’ capping enzymes), modified nucleotides (e.g. pseudouridine triphosphate used for the manufacturing of Moderna’s and Pfizer/BioNTech’s Covid-19 vaccine), cationic lipids used in the LNP formulations, plasmid DNA and T7 RNA polymerase enzymes [7,47]. The CapEx, OpEx, production scale and batch number ranges and values were presented in **Figure 4** on a 1 billion dose annual throughput basis, however, by linear extrapolation the CapEx, OpEx, production scale and batch number ranges and values can be approximated for producing vaccines for the global population. Therefore, the CapEx, OpEx, production scale and batch number values for meeting a c. 15 billion dose annual production target will be 15-fold higher than the values presented in **Figure 4**.

The fill-to-finish costs is additive to the DS production costs. By filling the AVV vaccine in 10-dose vials and by filling mRNA and saRNA vaccines in 5-dose vials, the production cost per dose, including both DS production and f2f, is 0.54, 2.39 and 0.39 USD per dose for AVV, mRNA and saRNA, respectively, cf. Table S1 and S2. For saRNA f2f cost per dose is the major cost contributor, for AVV the DS production and fill-to-finish costs are similar, whereas for mRNA the DS cost per dose accounts for most of the total production costs. The DS amount per vaccine dose could in principle decrease, not only for mRNA vaccines, but for all three platform technologies when developing second and third generation vaccines.

The global demand for COVID-19 vaccines is estimated at 15.6 billion doses, assuming a 2-dose regimen for the global population. The most optimistic estimate for current annual production capacity is 10 billion doses [48]. Therefore, there is a shortfall of at least 5.6 billion doses, without accounting for potential immunization demand caused by vaccine-escaping variants [49]. The resource requirements for meeting this shortfall computed based on the global sensitivity analysis results are illustrated in **Figure S5**. Therein, the violin plots show the ranges and probability distributions of the CapEx, OpEx, production scales and number of batches required to produce the 5.6 billion doses shortfall within a year using the three vaccine production platform technologies. For the risk analysis, worst-case scenarios can be defined at the maximum resource requirement values, illustrated by the maximum (100th percentile or Q4) top whisker. As shown by the probability distribution, there is a very small chance for this worse-case scenario to materialize based on this analysis. However, even in this worst-case scenario the benefits of establishing new production capacity based on all three platform technologies outweigh the costs by several orders of magnitude when considering the mortality, healthcare burden of the COVID-19 pandemic and economic decline. It is estimated that the pandemic has cost the global economy over 10 trillion USD [50], and the UN projects that the COVID-19 pandemic will reduce the global economy by a further 8.5 trillion USD over a 2-year period [51]. These substantial detrimental impacts of the COVID-19 pandemic can be avoided by comparatively small stimuli in the form of capital and operating costs, ranging between several hundred million to a few billion USD, as shown in **Figure S5A** and **S5B**, respectively. Based on these expenses the total drug substance production capacity shown in **Figure S5C** can be built to produce the number of batches (**Figure S5D**) required to meet the current shortfall. However, it is worth noting that such investments have to be made ideally in advance, or as soon as possible, considering the years-long timescale required to build such manufacturing capacity [1]. If this is built based on platform technologies, such as the RNA and AVV platforms, the resulting facilities could be used for producing a wide range of vaccines over their lifetime. Besides the financial resources, key raw materials (e.g. 5’ cap analogues or capping enzymes, cationic lipids and pseudouridine triphosphate), expertise and facilities for mass-producing mRNA vaccines and consumables for producing all vaccines are also in limited supply. An analysis of the material, consumables, labour and facility requirements for mass-producing mRNA vaccines for pandemic response has been carried out [47].

A large share of the COVID-19 vaccine shortfall described above is likely to be met by adapting or re-purposing manufacturing facilities that were used to manufacture other vaccines and biopharmaceuticals pre-pandemic. However, the healthcare impact of not sustaining routine childhood immunisations can outweigh that of the COVID-19 pandemic, especially in Africa [52,53]. Thus, it is crucial to manufacture and supply lifesaving vaccines against all vaccine-preventable diseases and to minimize the disruption in manufacturing of non-COVID-19 vaccines caused by the manufacturing of COVID-19 vaccines. This is even more important in the likely scenario of needing periodic, e.g. annual, COVID-19 booster doses to immunize adults at risk of severe COVID-19 and frontline workers in the foreseeable future [54]. If such one-dose booster vaccinations are to be required every year, building dedicated Covid-19 vaccine production facilities is a viable option. These facilities can be designed around platform technologies, enabling the production of vaccines or candidates against future SARS-CoV-2 variants, new coronavirus strains, or other emerging viruses [1,22]. The global population vulnerable to COVID-19, including people aged over 60 and adults with underlying medical conditions is around 2.2 billion [48]. In this scenario, ≈0.2 billion frontline personnel might also be required to be immunized to prevent the spread of the disease [48]. Therefore, in the single-dose boost scenario ≈2.4 billion COVID-19 vaccine doses would be required annually. The resource requirements for producing the annual boost vaccinations are illustrated in **Figure S6**. Therein, the violin plots show the ranges and probability distributions of the CapEx, OpEx, production scales and number of batches required to produce the 2.4 billion boost doses per year using the three vaccine production platform technologies.

Additionally, by investing in dedicated COVID-19 vaccine production facilities for supplying the annual COVID-19 boost vaccination the severe healthcare impact of other vaccine-preventable diseases can also be minimized by reinstating the production scale of other vaccines and biopharmaceuticals. Since vaccinations started in late 2020 or early 2021, administration of annual booster shots would potentially begin by the end of 2021. Using these platform, booster COVID-19 vaccines can be produced relatively quickly, as can production of new vaccines against emerging variants (which would still require clinical trials). The rate of mass-producing the 2.4 billion booster vaccinations will depend mostly on the platform technologies used, available production capacity and amount of drug substance per dose, which can be antigen specific. A feasible option would be to combine the annual COVID-19 booster dose with the annual influenza vaccine into a multivalent vaccine. For this, both vaccines can be produced using the same platform technologies. In this case, in contrast to current approaches, the influenza vaccine could be produced on demand for the strain in circulation, without the need of forecasting the 3-4 most prevalent influenza strains more than 6 months ahead of the start of the vaccination programme [55,56]. However, manufacturing the DS for multivalent vaccines comes with similar complexities and costs as a manufacturing of several monovalent vaccines. Since currently the manufacturing of vaccines represents the bottleneck for global immunization programs the need to develop and manufacture vaccines against a new variant would further limit immunization rates because more vaccine DS doses are required to immunize a person. Importantly, these platform technologies can speed up both the development of candidate vaccines and the manufacturing of regulatory-approved vaccines against a wide range of viral diseases, including currently known and currently unknown, future diseases.

## 3. Methodology

### 3.1. Vaccine production process modelling

The modeling of AVV, mRNA and saRNA drug substance production as well as drug product fill-to-finish was carried out using SuperPro Designer (Version 11, Build 2) by Intelligen, Inc. Further details are available in the SI document.

### 3.2. Data sources and assumptions

Information regarding mRNA and saRNA vaccine production processes and costs was obtained from the scientific literature [38,57–63], patent databases [26–29,64], from GMP grade material suppliers [31,65,66] and from experts [31,67,68]. Information regarding mRNA drug substance amount per dose was based on clinical trial databases [41,42,69,70] and the scientific literature [71]. For saRNA vaccines the drug substance amount per dose was obtained from the clinical trial registry [72]. Information on AVV vaccine production was obtained from the scientific literature [73,74] and consultation with experts [75]. The AVV vaccine production process was modelled based on the manufacturing of the replication-deficient chimpanzee adenovirus-vectored (ChAdOx1) vaccine which was co-developed by Oxford University and AstraZeneca plc. Information on AVV drug substance amount per dose was based on clinical trial databases [39,40,43,44,76–78]. Information on fill-to-finish technologies was obtained from the literature [15,79–81], equipment suppliers [14,16,82,83] and industry experts [84,85]. Additional production process data for all drug substance and drug product manufacturing processes were retreived from the equipment, materials, utilities and cost databases in SuperPro Designer [86]. The annualized CapEx is included in the OpEx. The CapEx value is also presented individually, in order to illustrate the financial requirements for building new facilities.

### 3.3. Sensitivity analysis

Variance-based stochastic global sensitivity analyses were conducted using SobolGSA Version 3.1.1 [87] under MatLab R2020a. 10,000 quasi-random scenarios were generated using Sobol sequences [23–25,88] according to the process parameter ranges and distributions in **Table 1** then passed to SuperPro Designer for evaluating the techno-economic KPIs in each scenario. A metamodel was generated in SobolGSA using the random-sampling high dimensional model representation (RS-HDMR) [89,90] based on which the main-effect and total-effect Sobol indices were derived [20]. A further 1,250 uncertainty scenarios were simulated in SuperPro Designer to test the predictions of the metamodel. The link between SobolGSA and SuperPro Designer was enabled by a Component Object Model (COM) interface in MS-Excel VBA available from from MS-Office 365 Enterprise. Further details are available in the SI document. Data processing and visualization/plotting is also described in the SI document.

#### Data Availability

Data is available from: https://github.com/ZKis-ZK/RNA_AVV_vaccine_production-cost_modelling_global_sensitivity_analysis

## 4. Conclusions

In this computational modelling study, the COVID-19 pandemic-response manufacturing performance of the AVV, mRNA and saRNA vaccine platforms has been assessed using techno-economic modelling and variance-based global sensitivity analysis. The three vaccine production processes have been presented and key design and operation uncertainties and variations were assessed in terms of their impact on the productivity and resource-intensity performance indicators of these processes. It was established that variations in both the annual productivity and cost per dose of AVV vaccines can be primarily attributed to variations in the scale and titre/yield of the production process. On the other hand, the variations in the annual productivity and cost per dose of mRNA and saRNA vaccines can be primarily attributed to variations in the RNA amount per dose. The saRNA platform is likely to be the fastest to meet the global demand, followed by the AVV and then by the mRNA platform. The results of these assessments depend on the specific characteristics, such as the production scale, at which these platform technologies are implemented, and these characteristics are fundamentally different among these platform technologies alongside the drug substance amount per dose and the time required to produce batches. The performance of the AVV platform can be improved by increasing the yield in the production bioreactor. Decreasing the RNA amount per dose, would improve the production rates and volumes of mRNA and saRNA vaccines and by increasing their (thermo)stability would improve their usability across the globe, including in low- and middle-income countries. To meet the current global projected vaccine shortfall of 5.6 billion doses and annual booster vaccination production, investments ranging from hundreds of millions to a few billion USD would be required, which is very small in comparison to the mortality, healthcare and economic cost of the COVID-19 pandemic, estimated at over 10 trillion USD. Overall, this model-based assessment can inform policymakers and vaccine manufacturers for risk assessment, on how to improve manufacturing and asset utilisation against COVID-19 and its variants, but also against future outbreaks due to the disease-agnostic nature of these vaccine production platforms. The platform technology-based dedicated COVID-19 vaccine production would prevent the reduction of the manufacturing throughput of other, non-COVID-19 vaccine and therapeutics, and would allow rapid-response vaccine production against a wide a range of viral targets. These platform technologies will enable faster vaccine development and production for overcoming future epidemics and pandemics, especially if surge manufacturing capacity is maintained during times without outbreaks.

## Acknowledgments

This research is partly funded by UK Research and Innovation (UKRI) via the Engineering and Physical Sciences Research Council (EPSRC) grant on COVID-19/SARS-CoV-2 vaccine manufacturing and supply-chain optimisation (EP/V01479X/1) and the Future Vaccine Manufacturing Research Hub at UCL-Oxford (EP/R013756/1). Additional financial support from the Department of Health and Social Care using UK Aid funding as managed by the EPSRC (EP/R013764/1) is also gratefully acknowledged. The views expressed in this publication are those of the author(s) and not necessarily those of the Department of Health and Social Care. The authors gratefully acknowledge insightful discussions with Robin Shattock (Imperial College London, UK), Sandy Douglas (The Jenner Institute, UK), Harvey Branton (Centre for Process Innovation, UK) and John Liddell (Centre for Process Innovation, UK).

## Author Contribution

Conceptualization: ZK, KT, BC, MMP, CK, NS; Methodology: ZK, KT, DI, BC, MMP, CK, NS; Software: ZK, KT, BC; Writing - Original Draft: ZK, KT; Writing - Review & Editing: ZK, KT, BC, MMP, CK, NS; Visualization: ZK, KT; Supervision: BC, MMP, CK, NS; Funding acquisition: BC, MMP, CK, NS.

