## Supplementary Information for "Pandemic-response adenoviral vector and RNA vaccine manufacturing"

Zoltán Kis*¥1, Kyungjae Tak*1, Dauda Ibrahim1, Maria M Papathanasiou1, Benoît Chachuat1, Nilay Shah1 and Cleo Kontoravdi¥1

*These authors contributed equally and share first authorship

#### Supplementary Results

##### 1.1. Background on vaccine drug substance production technologies presented in this study

There are several vaccine drug substance (DS) production technologies in use and in development around the world to produce vaccines against SARS-CoV-2 [1–3]. Outside of China, vaccine production platform technologies, such as the mRNA and adenovirus vector vaccine platforms, were the fastest to produce vaccines which have gained emergency use authorisation from the regulatory authorities to manufacture vaccines against SARS-CoV-2 [1–5]. In China, conventional inactivated whole viral vaccine production technologies are also being used to produce vaccines and candidate vaccines [1–3]. The adenoviral vector vaccine platform uses mammalian cells, such as human embryonic kidney 293 (HEK293) cells, to express the adenoviral vectors [6–9]. The RNA platforms utilize a cell-free enzymatic reaction based on the T7 RNA polymerase enzyme to synthesize the RNA polymer of the vaccine [10–12]. This RNA molecule is then encapsulated into LNPs to prevent degradation and aid the delivery of the RNA into the cells of the human body [10–12]. The whole viral vaccine is produced using Vero cells and it is inactivated with formaldehyde [2,13]. Recombinant protein subunit vaccine candidates are produced using the insect cell – baculovirus expression system [1,2,14–16]. Most of the COVID-19 vaccines are produced in facilities which before the COVID-19 pandemic were used to produce other vaccines and biopharmaceuticals. Thus, the production of these non-COVID-19 vaccines and biopharmaceuticals is disrupted by COVID-19 vaccine production.

Once fully developed and validated, vaccine production platform technologies will be more suitable than conventional vaccine production technologies to rapidly deploy vaccines at high volumes against a new pathogen that causes a global pandemic [10]. That is because in principle any viral vaccine candidate against any known or currently unknown viral disease can be produced with the same production process, same type of raw materials and consumables, same formulation components, same fill-to-finish processes, using the same quality control approach and the same personnel. This would reduce process development and optimisation timelines, the need for construction of production facilities, to set up quality control methodologies and to train personnel when switching to produce a new vaccine against a new viral target [10].

In order to aid vaccine manufacturing and to inform policymakers we present our techno-economic modelling results for the three promising vaccine production technologies: (1) the adenovirus-vectored vaccine (AVV) platform, based on the chimpanzee replication-deficient adenovirus-vectored vaccine (ChAdOx1), (2) the messenger RNA (mRNA) vaccine formulated in lipid nanoparticles (LNPs), and (3) the self-amplifying RNA (saRNA) vaccine formulated in lipid nanoparticles (LNPs) [1–12]. All three DS platform technologies produce genetic instructions based on which the cells of the human body produces the antigen of the vaccines, which in case of COVID-19 vaccines is the spike protein of the SARS-CoV-2 virus [1–12].

##### 1.2. Chimpanzee adenovirus-vectored (ChAdOx1) vaccine drug substance production process modelling

The AVV production process was modelled based on the manufacturing of the replication-deficient chimpanzee adenovirus-vectored (ChAdOx1) vaccine which was co-developed by Oxford University and AstraZeneca plc. The ChAdOx1 production process starts with preparing the HEK293 cell seed train and the adenovirus inoculum seed train. For this, the HEK293 cells are cultured at increasing volumes until the culture amounts required for the production bioreactor (commonly at 2000 L working volume) scale are obtained. In the production bioreactor the HEK293 cells are first cultured to reach the required cell densities and then these cells in which the adenovirus can replicate are infected to produce the adenovirus. This adenovirus has been genetically modified to express the SARS-CoV-2 spike protein and it cannot replicate in human cells, thus it is a non-replicating vector for delivering the genetic material to express the SARS-CoV-2 spike protein. The cell culture and virus production in the bioreactor takes around 55-60 days and it can be operated in fed-batch mode. The production titre was assumed at $5 \times 10^{14}$ viruses per L of culture in the bioreactor. From the bioreactor, the solution enters the downstream separation and purification section of the process whereby cells are initially lysed then the larger impurities (such as cell debris) are removed using microfiltration where the adenoviral vectors flow through the filter. Next, another particle size-based separation is performed, where the tangential flow ultrafiltration/diafiltration is carried out to retain adenoviruses by the filter. Here the adenoviruses are washed, and the buffer is replaced with a buffer suitable for the subsequent ion-exchange chromatography step. In the ion-exchange chromatography step the adenoviral vectors are separated based on electrical charge differences that exist between these adenoviral vectors and other impurities. Following elution from the ion-exchange chromatography, the adenoviral vector solution is sterile filtered, and the buffer can be exchanged for the formulation buffer, using tangential flow ultrafiltration/diafiltration. The total losses in the downstream purification process are around 50%. The production process is assumed to run 335 days per year. After the adenovirus vaccine DS (active ingredient) is produced, it is then formulated and filled into vials or other containers, often at a different facility at a different location. The number of viral particles per vaccine dose was set to $5 \times 10^{10}$. The SuperPro Designer files for the ChAdOx1 vaccine DS production process is available at https://github.com/ZKis-ZK/RNA_AVV_vaccine_production-cost_modelling_global_sensitivity_analysis.

##### 1.3. Messenger RNA (mRNA) and self-amplifying RNA (saRNA) vaccine drug substance production process modelling

Both the mRNA and saRNA DS is produced using the same production process. The difference is that for the mRNA the amount per dose is 100 µg whereas for saRNA it is 1 µg. The mRNA and saRNA vaccine production process starts with a cell-free enzymatic reaction, whereby the mRNA or saRNA molecule is synthesized using the T7 RNA polymerase enzyme based on a DNA template. The mRNA or saRNA synthesis, ( in vitro transcription) reaction takes 2 hours and then the DNA template is digested using the DNAse I enzyme in 15 minutes and then the solution enters the downstream purification section. This reaction mix has a well-defined and simple composition compared to cell-based culture broths, thus the downstream purification is also relatively simple. Downstream purification can consist of tangential flow filtration, whereby the mRNA or saRNA which is the largest size component is retained by the filter and the other smaller size components flow through the filter. Next, the mRNA or saRNA can be further purified using Capto Core 700 multi-modal chromatography. After this the buffer is exchanged for the formulation buffer using tangential flow ultrafiltration/diafiltration. Next, formulation in LNPs takes place, which is the longest duration operation, hence the bottleneck, taking around 9-10 hours to complete. The formulation commonly takes place at the DS manufacturing site, to stabilise the RNA molecule. Following formulation, the solution is sterile filtered and can be shipped to the fill-to-finish site. The losses in the downstream section of the mRNA and saRNA production process were assumed at 30% and it was assumed that an additional 20% losses occur in the LNP formulation process. This way, the total losses in the formulated DS production process, from the beginning of the downstream separation and purification section till the end of the formulation section were considered 44% in the baseline scenario. The SuperPro Designer files for the mRNA and saRNA vaccine DS production process is available at https://github.com/ZKis-ZK/RNA_AVV_vaccine_production-cost_modelling_global_sensitivity_analysis.

#### 1.4. Fill-to-finish processes modelling

The following three different fill-to-finish processes have been modelled: conventional fill-to-finish in 10-dose glass vials, blow-fill-seal in single-dose ampules and the new 200-dose bag Intact™ Modular Filler [10,17]. All fill-to-finish processes were modelled to operate in continuous mode with multiple runs for a maximum of 330 days per year. All fill-to-finish process models consist of two parts: 1) solution preparation section where the required formulation or dilution operations take place, and 2) the filling section where filling into vials or other containers, capping (if required), inspection, labelling and packaging takes place with required transfer steps in between these unit operations. The capping operation is only required in the conventional fill-to-finish process. The cycle time slack (time between subsequent production runs) was set to 2 hours [18]. The product failure rate was set to 5% in all fill-to-finish process models.

Conventional filling into 10-dose glass vials was modelled based on large-scale filling lines capable of filling 400 vials/minute, such as the Bosch MLF series [19,20]. The annual equipment utilization rate ( overall equipment effectiveness, OEE) was considered at 62% [21–23]. The model in SuperPro designer was built using the database of equipment and unit operations and based on information from suppliers [18,21,23]. The purchase price of the empty 10-dose glass vial was considered at $2.2 per vial [22].

For blow-fill-seal a the process was modelled based on filling equipment such as the BP460-20 BFS Aseptic Filling Machine [24,25]. This machine can fill ampoules with volumes between 0,1–20 mL with a filling rate of up to 34000 ampoules per hour [24,25]. Based on information from the equipment supplier, the purchase price of the filling machine was assumed at 7 million USD, the purchase price of the inspection system was 2 million USD and the secondary packaging equipment purchase price was 1.5 million USD [21]. The equipment has a size of 5 x 2.9 x 4.3 m (L x W x H) and requires a room size of 7.5 x 6 x 5 m (L x W x H) [24,25]. The equipment requires a Grade C or ISO class 8 room for operation, as the machine generates its own Grade A or ISO class 5 clean environment, similarly to a laminar flow isolator [21,24]. The annual equipment utilization rate (

overall equipment effectiveness, OEE) was considered at 85% [21]. The cost of the material for generating the empty single-dose 0.5 mL ampoule was considered at $0.002 per dose [21]. Based on this information, a production process model has been built in SuperPro Designer. Filling into single-dose 0.5 mL ampoules was modelled at a rate of 550 ampoules/minute (corresponding to 33000 ampoules per hour) for the baseline scenario. Given the relatively small footprint of the equipment, 3 blow-fill-seal machines were modelled to operate in parallel in stagger mode to increase productivity and de-bottleneck the production process.

For modelling the 200-dose bag Intact™ Modular Filler, one filling machine with 15 parallel filling needles has been considered [17]. This machine is capable of filling 116250 pouches per day, considering 3 shifts per day [17,26]. This corresponds to 23 million doses per day when filling into 200-dose pouches (bags) [17,26]. The annual equipment utilization rate (overall equipment effectiveness, OEE) was modelled at 87% [17,26]. The equipment requires a Grade C or ISO class 8 room for operation, as the machine generates its own Grade A or ISO class 5 clean environment, similarly to a laminar flow isolator [26]. The purchase price of the filling line with 15 filling needles is 12 million USD [26]. The purchase price of the inspection system was 2 million USD and the secondary packaging equipment purchase price was 1.5 million USD. The purchase price of the empty 200-dose pouch was $4.03 [26]. This information has been compiled into the SuperPro Designer process model. The Intact™ Modular Filler can also fill into 400-dose bags, doubling the productivity of the system. In addition, a bespoke multi-dose and multi-use syringe is also available for this system which can substantially reduce administration costs [17,26]. Filling into 400-dose bags and using the multi-use syringe has not been modelled in this study.

The assumptions, input variables and key performance indicator outputs for these fill-to-finish technologies are summarized below in **Table S1**. Lyophilized vaccine production processes were not included in this study due to the fast use and short-term storage of vaccines due to high and urgent demand of pandemic-response vaccines and the additional times required for developing a lyophilised formulation. As expected, filling into larger multi-dose vials increases the production throughput in terms of doses filled per unit time compared to filling into smaller multi-dose or single-dose vials. The 200-dose bag filling technology is best suited for pandemic response vaccine production, if the stability of the vaccine allows. The cost per dose is also decreasing as the container size increases, however using blow-fill-seal a low cost per dose can be achieved when filling into smaller (i.e. single-dose) ampules as well.

**Table S1**. Model inputs and outputs for fill-to-finish technologies obtained using SuperPro Designer.

| Parameter name | Unit | Conventional liquid fill in 10-dose vials | Conventional liquid fill in 5-dose vials | Blow-fill-seal in single dose*** | 200-dose bag filling |
|---|---|---|---|---|---|
| Annual production amounts* | vials/year | $1.1 \times 10^8$ | $1.4 \times 10^8$ | $5.8 \times 10^8$ | $3.1 \times 10^7$ |
| Number of doses per container | dose/vial | 10 | 5 | 1 | 200 |
| Annual production amounts | doses/year | $1.1 \times 10^9$ | $7.0 \times 10^8$ | $5.8 \times 10^8$ | $6.1 \times 10^9$ |
| Time for 1 billion doses | years/fill line | 0.85 | 1.43 | 1.73 | 0.16 |
| Number of batches per year | batches/year | 363 | 222 | 502 | 168 |
| Amount per batch | Vials/batch | $3.1 \times 10^5$ | $6.3 \times 10^5$ | $1.1 \times 10^6$ | $1.8 \times 10^5$ |
| Capitals expenses (CapEx) | USD/facility | $1.8 \times 10^8$ | $1.8 \times 10^8$ | $3.8 \times 10^8$ | $2 \times 10^8$ |
| Operating expenses (OpEx) | USD/year | $3.1 \times 10^8$ | $2.6 \times 10^8$ | $8.4 \times 10^7$ | $1.6 \times 10^8$ |
| Cost per dose** | USD/dose | 0.27 | 0.37 | 0.15 | 0.027 |

* Represent the annual filling capacity, taking into account the overall equipment effectiveness.
** Time required to fill 1 billion doses per production line.
*** Includes all the costs associated with drug product fill-to-finish calculated using SuperPro Designer, excluding the DS costs.
*** Based on 3 fillers operating in stagger mode for process de-bottlenecking and efficiency increase, for detail see SI.

The overall production bottleneck, when considering both DS production and f2f, depends on the combination of the specific technologies. For example, in case of filling AVV vaccines in 10-dose vials, the bottleneck will be in the DS production when one AVV DS production line with baseline characteristics (cf. **Table S2**) at the common 2000 L bioreactor working volume scale is coupled to a single 10-dose vial filling line which fills at 400 doses per minute at 60% overall equipment effectiveness (OEE). However, depending on the uncertainty realization (e.g. when larger DS production scales are also considered), the DS production rate might surpass the fill-to-finish rate, as shown by the global sensitivity analysis results presented in **Figure 3A** (in the main text) compared to the values shown in **Table S1**. Specifically, the time required to produce 1 billion AVV DS doses is $0.96^{+0.76}_{-0.39}$ years (median of 0.96 years, the 75th minus 50th percentile of 0.76 years, and 25th minus 50th percentile of -0.39 years) and filling 1 billion doses would require 0.85 years per production line. If mRNA vaccines are filled into 5-dose vials (e.g. BioNTech/Pfizer vaccine) at a rate of 400 vials per minute, the overall production bottleneck is at the DS production stage, when comparing the $2.19^{+1.91}_{-0.9}$ years per production line value from **Figure 3A** with the 1.43 years per filling line value from **Table S1**. The baseline mRNA vaccine production rate (cf. **Table S2**) is also slower then filling into 5-dose vials. If an mRNA vaccine production line is coupled with a 10-dose vial (Moderna vaccine) filling line which fills at 400 vials per minute, the overall production bottleneck would again be at the DS production stage. On the other hand, in case of saRNA DS vaccine production coupled with f2f into 5-dose vials at the 400 vials/minute rate, the overall production bottleneck is at the f2f stage. This is the case regarding both the saRNA baseline annual productivity (**Table S2**) and the $0.083^{+0.084}_{-0.041}$ years per saRNA production line value obatined from the global sensitivity analysis (**Figure 3A**). This fill-to-finish bottleneck for saRNA vaccine production can be removed by using a higher throughput fill-to-finish technology, such as the INTACT™ Modular Filler, if the (thermo)stability of the saRNA vaccine is compatible with this technology [17,26]. To this end, besides filling in 200-dose bags, a highly thermostable vaccine can in principle also be filled into 400-dose bags at a rate of around 1.24 billion doses per month per filling line based on filling 116250 pouches per day in 3 work shifts using a 15-needle INTACT™ Modular Filler from MEDInstill Development LLC, operating at 90% overall equipment effectiveness (OEE) which translates to 27 days of operation per 30-day calendar month [17,26]. On the other extreme, if vaccines need to be lyophilized and filled into single-dose vials, only 4.36 million doses can be filled per month per facility assuming 60% overall equipment effectiveness (OEE). However, vaccine lyophilization seems unlikely for an emergency-use pandemic-response vaccine as the storage time will be short, and high-volume and rapid production is required.

The thermostability of the vaccines can also substantially impact the supply chain and administration of vaccines. This way, in a worst-case scenario, if vaccines need to be stored at -80°C throughout the transportation and storage step, distribution logistic difficulties and costs will increase. Delivery of vaccines at -80°C is extremely challenging or even impossible to rural areas of developing countries with tropical and sub-tropical climate, where road and/or electricity infrastructure is inadequate, such as large parts of Sub-Saharan Africa. Administration of a lyophilized vaccine would also involve additional dissolution/resuspension steps, which would take precious medical-staff time, which will be on extremely high demand during pandemic-response vaccination campaigns. The number of vaccine doses require per person would also impact the required production amounts and the supply-administration chain.

#### 1.5. Comparative technological assessment of COVID-19 vaccine production platforms

The main process modelling assumptions and KPIs are listed in **Table S2**. This table contains information pertaining to the baseline scenario, which at the time of this study contains the most up-to-date information available to the authors.

As shown in **Table S2**, the mRNA and saRNA processes are normally implemented at scales which are two and three orders of magnitude smaller, respectively, compared to the AVV platform. In addition, the mRNA and saRNA production processes are also substantially faster with batch durations in the order of approximately 2 days compared to the 2-months batch duration of the AVV platform. As a consequence, the productivity of the mRNA and saRNA platform can be two and four orders of magnitude higher than the AVV platform, respectively, when expressed in number of doses per L of bioreactor working volume per unit time. These results show that the mRNA and especially the saRNA platform are better suited for global pandemic-response vaccine production compared to the AVV platform. The AVV vaccine production is based on mammalian cells, requiring to maintain cell viability and optimal cell functions (including metabolism, gene expression regulation, cell signalling, etc.) during the two-months long cell culture phase, thus AVV vaccine production is associated with a high level of inherent biological variability. On the other hand, RNA vaccines are produced in a much faster cell-free enzymatic process, where the complexities and variabilities associated with mammalian cell culture are absent. This makes the RNA vaccine production process simpler, more robust and reproducible from batch to batch.

**Table S2.** Baseline scenario assumptions, modelling inputs and key performance indicators for the AVV, mRNA and saRNA vaccine platform technologies obtained using SuperPro Designer.

| Parameter name and unit | AVV platform | mRNA platform | saRNA platform |
|---|---|---|---|
| Production scale – Bioreactor working volume [L] | 2000 | 30 | 5 |
| Production titre in bioreactor | $2.5 \times 10^{14}$ vir.×L$^{-1}$ | 5 g×L$^{-1}$ | 5 g×L$^{-1}$ |
| Total losses in downstream purification [%] | 50 | 44* | 44* |
| Duration of a batch [hours × batch$^{-1}$] | 1,440 | 48* | 42* |
| Cycle Time, overlapping batches [hours × subsequent batches$^{-1}$] | 150 | 18* | 17* |
| Drug substance amount per dose | $5 \times 10^{10}$ vir.×dose$^{-1}$ | 100 µg×dose$^{-1}$ | 1 µg×dose$^{-1}$ |
| Doses per bioreactor working volume [doses × L$^{-1}$ × day$^{-1}$]** | 315 | $3.5 \times 10^4$ | $3.6 \times 10^6$ |
| Doses per batch | $4.7 \times 10^6$ | $7.8 \times 10^5$ | $1.3 \times 10^7$ |
| Maximum number of batches per year | 45 | 444 | 470 |
| Maximum doses per year | $2.1 \times 10^8$ | $3.5 \times 10^8$ | $6.1 \times 10^9$ |
| Capital costs, including working capital [USD] | $2.2 \times 10^8$ | $7.7 \times 10^7$ | $2.4 \times 10^7$ |
| Operating Costs [USD × year$^{-1}$]*** | $5.8 \times 10^7$ | $7.0 \times 10^8$ | $1.2 \times 10^8$ |
| Drug substance Cost per dose [USD × dose$^{-1}$] | 0.27 | 2.0 | 0.0160.02 |

* includes the LNP formulation step, because this preferentially occurs at the site of the RNA DS production. This LNP formulation step can be the longest duration procedure for RNA DS production.

** Doses per L of bioreactor working volume per day, accounting for the losses in the downstream purification and assuming 335 working production days per year.

*** The operating costs (OpEx) include the annualised capital costs (CapEx), calculated using the straight-line depreciation method. The CapEx was included separately from the annualized OpEx to describe the financial resources required to construct facilities.

Low capital investment cost is indicative of lower footprint manufacturing facilities which can be constructed faster and can also be scaled out more rapidly. The process development and construction of these small-scale manufacturing processes can be further accelerated when single-use production equipment is used, however procurement of single-use equipment currently (from late 2020 until mid 2021) represents a major bottleneck.

Since these are new vaccine platform technologies deployed for emergency use against a previously unknown disease, these production platforms are surrounded by numerous uncertainties. Understanding the impact of these uncertainties on the performance of the production process will also support platform re-utilization against future viral pathogens [10]. For this purpose, we have carried out sensitivity analysis, in which the sensitivity of the KPIS of annual production amounts and cost per dose to variations in the model inputs of production scale, titre and DS amount per dose was computed. All relevant model inputs were evaluated, and the ones included in **Figure S1** were found to have the highest impact on the model output KPIs. The DS amount per dose has been determined for those vaccines which have been granted emergency use authorisation by the regulatory authorities. However, the DS amount per dose can vary in case of future vaccines produced against new diseases using the same platform technologies, also depending on the immunogenic properties of the antigen. There is also new evidence indicating that the amount per dose of the BioNTech mRNA vaccine could be reduced [27]. By including baseline scenarios for the high-dose mRNA vaccine (based on Moderna's COVID-19 vaccine) and a low dose saRNA, the characteristics of the RNA platform are better represented from a production performance point of view. Based on the modelling results, as shown in **Figure S1**, the annual production amount is proportional to the scale of the production process and production titres, however, it is inversely proportional to the amount per dose. On the other hand, the cost of the DS per dose shows an abrupt non-linear drop as the scale and titre increases and then the cost per dose flattens out as the input variables further increase. The DS cost per dose has a linear relationship with the amount per dose. The capital costs (CapEx) and operating costs (OpEx) change linearly in function of scale of the production process but are not impacted by changes in production titres and DS amounts per dose. These observations and trends hold true for all of the 3 platform technologies, as shown in **Figure S1**. The values indicated by grey dots and grey triangles correspond to the baseline scenarios illustrated in **Table S2**. These plots also indicate the potential for cost reduction and productivity increase in function of production scale and titre increase and DS amount per dose decrease, when only one factor is changed at a time.

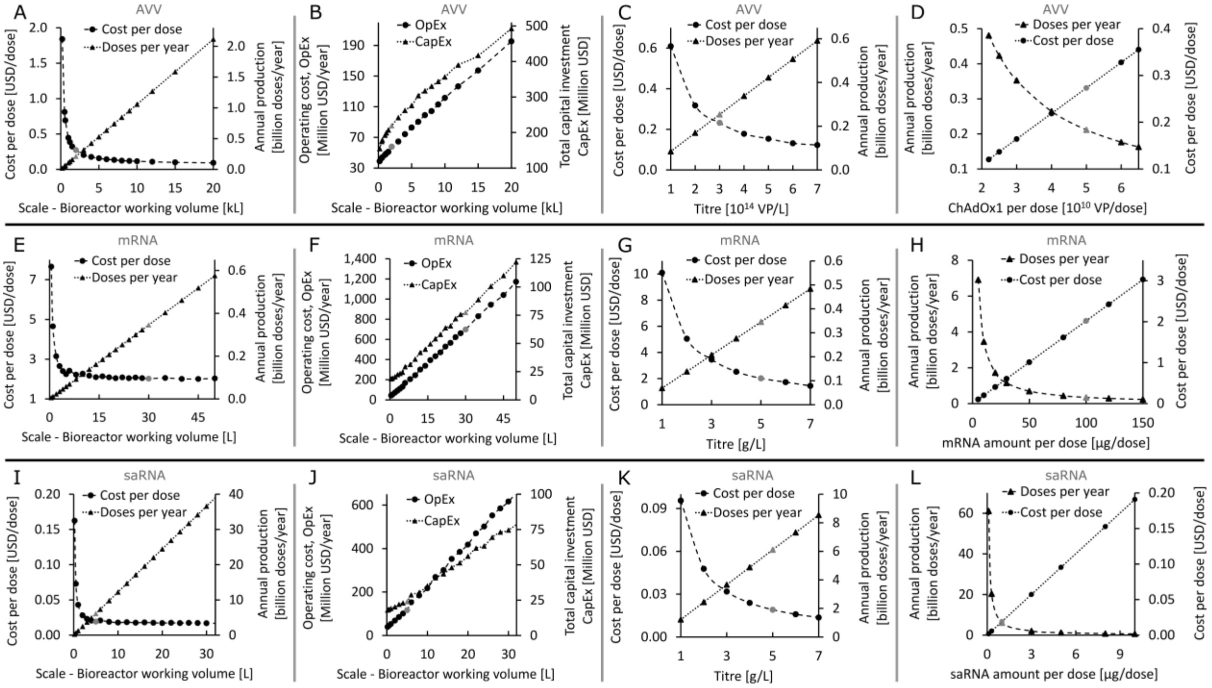


**Figure S1**. The individual impact of AVV, mRNA and saRNA vaccine DS production scale, titre and DS amount per dose on the annual production amounts and production costs. A-D (top row), E-H (middle row), I-L (bottom row) shows AVV, mRNA and saRNA vaccine DS production performance, respectively. The values indicated by grey dots and grey triangles correspond to the baseline scenarios. **A.** The impact of production process scale, represented by bioreactor working volume in thousands of L, on the AVV DS cost per dose and annual production amounts. **B**. The effect of AVV production process scale indicated by the bioreactor working volume on OpEx and CapEx. **C**. The dependence of the AVV DS cost per dose and annual production amount on the production titre expressed in VP per litre at harvest in the production bioreactor. **D**. The influence of the AVV amount per dose on the annual production amounts and cost per dose. **E.** The impact of production process scale, represented by bioreactor working volume in L, on the mRNA DS cost per dose and annual production amounts. **F**. The effect of mRNA production process scale indicated by the bioreactor working volume on OpEx and CapEx. **G**. The dependence of the mRNA DS cost per dose and annual production amount on the production titre expressed in g per litre at the end of the enzymatic bioreaction. **H**. The influence of the mRNA amount per dose on the annual production amounts and cost per dose. **I.** The impact of production process scale, represented by bioreactor working volume in L, on the saRNA DS cost per dose and annual production amounts. **J**. The effect of saRNA production process scale indicated by the bioreactor working volume on OpEx and CapEx. **K**. The dependence of the saRNA DS cost per dose and annual production amount on the production titre expressed in g per litre at the end of the enzymatic bioreaction. **L**. The influence of the saRNA amount per dose on the annual production amounts and cost per dose.

In case of AVV vaccine production, the major cost component is the depreciation and facility dependent cost, as shown below in **Figure S2**. On the other hand, for mRNA and saRNA vaccine production, the major operating cost component is the material costs, out of which the cost of the 5' cap analogue (e.g. CleanCap AG and CleanCap AU from TriLink BioTechnologies, Inc. [28–30]) is the major cost driver,, for further details see **Figure S2** in the Supplementary Information document.

A

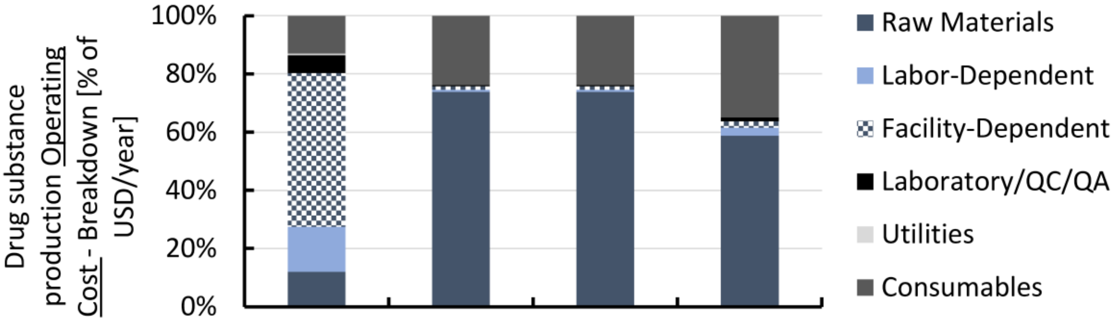


| Platform | AVV | mRNA | mRNA | saRNA | |
|---|---|---|---|---|---|
| Amount per dose | 5 x $10^{10}$ vir./dose | 100 µg/dose | 30 µg/dose | 1 µg/dose | |
| UTP | modified | modified | modified | Wild-type | |
| Common Scale [L] | 2000 L bior. w. v. | 30 L bior. w. v. | 30 L bior. w. v. | 7 L bior. w. v. | L of bioreactor working volume |
| OpEx [$/Year] | 57.79 Million | 697.97 Million | 697.97 Million | 168.46 Million | Total drug substance production Operating cost – Baseline Scenario [$/year] |

B

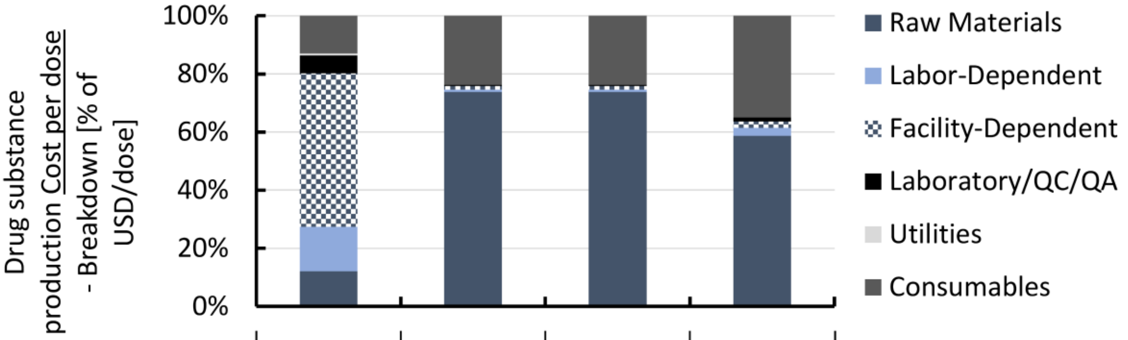


| Platform | AVV | mRNA | mRNA | saRNA | |
|---|---|---|---|---|---|
| Amount per dose | 5 x $10^{10}$ vir./dose | 100 µg/dose | 30 µg/dose | 1 µg/dose | |
| UTP | N.A. | modified | Modified | Wild-type | |
| Common Scale [L] | 2000 L bior. w. v. | 30 L bior. w. v. | 30 L bior. w. v. | 7 L bior. w. v. | L of bioreactor working volume |
| Doses per year | 211.48 6Million | 345.09 Million | 1.15 Billion | 8.55 Billion | Doses produced per year per facility |
| Cost per dose | 0.27 | 2.02 | 0.61 | 0.02 | Total drug substance production cost per dose – Baseline Scenario [$/dose] |

**Figure S2**. Breakdown of DS production annual operating costs and cost per dose for AVV and the LNP-formulated RNA vaccine types. **A**. Share of operating cost (OpEx) components. The percentage of each OpEx component is shown on the y-axis and the four vaccine types are shown on the x-axis. The table below the x-axis also indicates the total OpEx in USD per year for a single facility with a single production line at the common scale. **B**. Share of cost per dose components. The percentage of each cost per dose component is shown on the y-axis and the four vaccine types are shown on the x-axis. The table below the x-axis also indicates the total cost per dose for the DS production for the four vaccine types. The number of DS doses produced per year are also shown for a facility at the common scale which is indicated in the table below the x-axis.

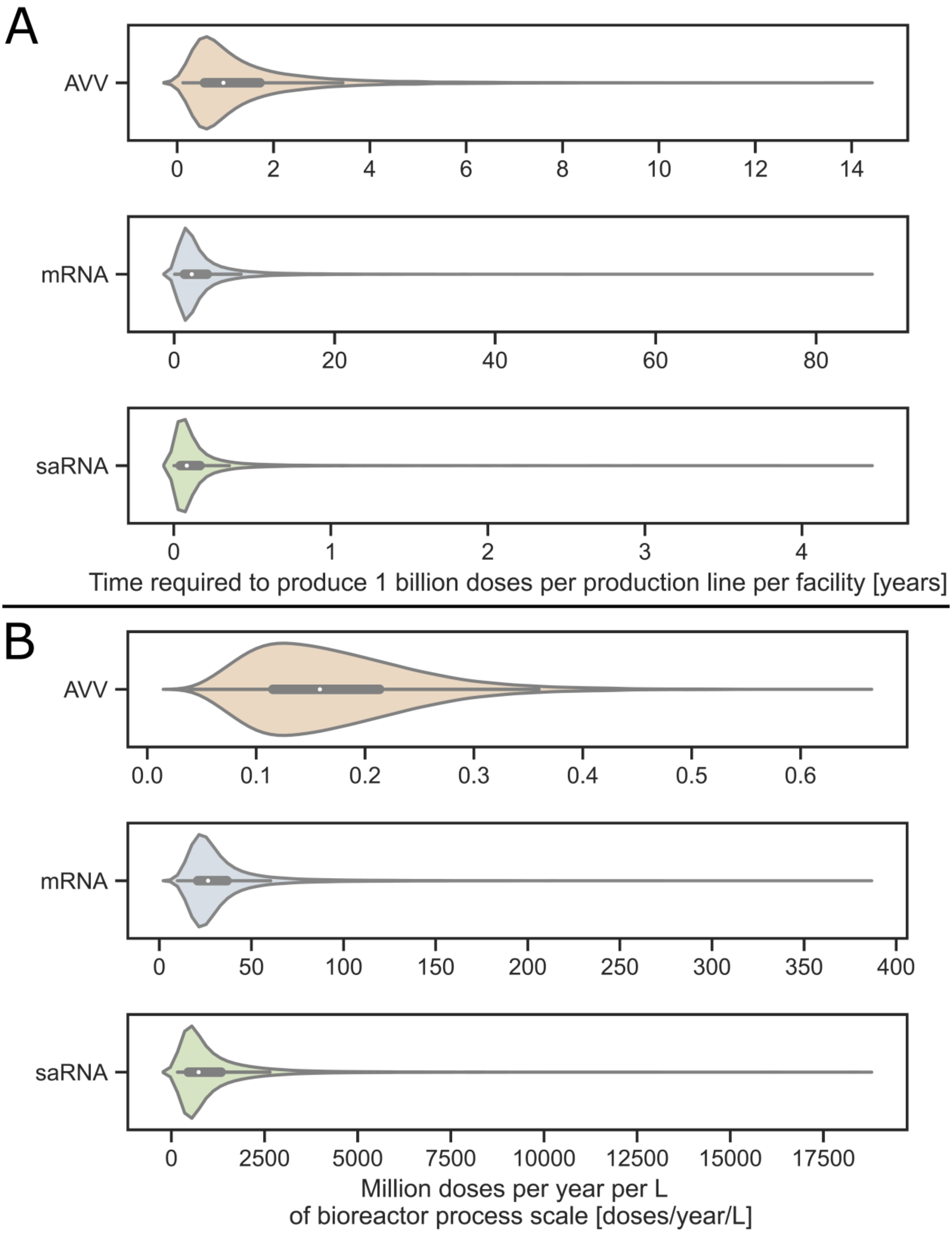


**Figure S3**. Speed and productivity of the AVV, mRNA and saRNA vaccine production platform technologies. Time requirements for producing DS for 1 billion COVID-19 vaccine doses using the AVV, mRNA and saRNA production platforms. A. The number of batches completed per year and the duration of a batch for DS production at the characteristic production process scale. **A**. Violin plots showing the computed time requirements for producing 1 billion AVV, mRNA and saRNA vaccine DS doses. The time is shown on the x-axis and the production technologies together are listed on the y-axis. **B**. Violin plots showing the number of vaccine doses produced per year per unit of process scale. The unit of process scale is expressed per L of bioreactor working volume. Box and whisker plots are shown in the centre of the all violin plots. The box and whisker plots show the minimum and maximum values, except outliers, with the ends of the whiskers; the 25th and 75th percentiles with the top and bottom of the boxes; and the median in the central part of the box. These global sensitivity analysis results were obtained based on the modelling inputs from Table 1 in the main text. This figure is equivalent to Figure 3 from the main text, however in Figure S3 all the data is included, whereas in Figure 3 the top 5% and bottom 5% of the values are excluded.

#### 1.6. Production process scales and resources required to produce multi-billion doses of Covid-19 vaccine

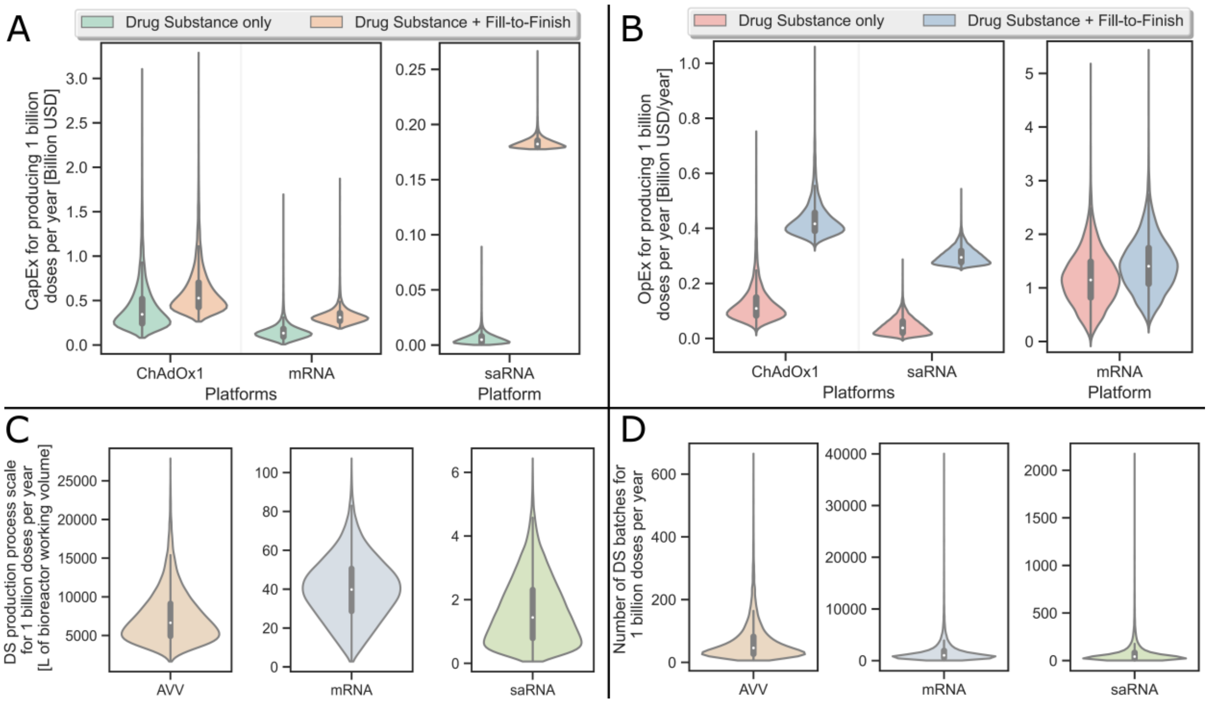


**Figure S4**. Violin plots showing the global sensitivity analysis of the resource requirements for producing 1 billion COVID-19 vaccine doses per year using the AVV, mRNA and saRNA production platforms combined with conventional liquid fill-to-finish. The inputs and their ranges used for this global sensitivity analysis are shown in Table 1 in the main text. In the centre of the violin plots, box and whisker plots are shown with the median values indicated by the white dots. **A.** Operating costs (OpEx) required to produce 1 billion doses per year of AVV, mRNA and saRNA vaccine DS and drug product. It was assumed that AVV vaccine is filled into 10-dose vials, whereas the mRNA and saRNA vaccine is filled into 5-dose vials. **B.** Capital costs (CapEx) required to produce 1 billion doses per year of the vaccine DS and drug product using the three platform technologies. AVV vaccine fill-to-finish was modelled based on 10-dose vials, whereas the mRNA and saRNA vaccine fill-to-finish was modelled based on 5-dose vials. **C**. Production process scales required to produce 1 billion doses of DS per year using the AVV, mRNA and saRNA vaccine production platforms. The scale of the production process is represented by the working volume in the bioreactor and the entire process is scaled based on the mass balances proportionally to the bioreactor working volume. **D**. Number of batches required to produce 1 billion doses of DS per year using the AVV, mRNA and saRNA vaccine production platforms. The box and whisker plots show the minimum and maximum values, except outliers, with the ends of the whiskers; the 25$^{th}$ and 75$^{th}$ percentiles with the top and bottom of the boxes; and the median in the central part of the box. This figure is equivalent to Figure 4 from the main text, however in Figure S4 all the data is included, whereas in Figure 4 the top 5% and bottom 5% of the values as well as the fill-to-finish values are excluded.

The violin plots in **Figure S5** show the ranges and probability distributions of the CapEx, OpEx, production scales and number of batches required to produce the 5.6 billion doses shortfall within a year using the 3 vaccine production platform technologies. The ranges and probability distributions of these resource requirements can be used for risk analysis. For this, inside the violin plots, box-and-whisker plots also show the minimum (0th percentile or Q0) and maximum (100th percentile or Q4) values using the extremities of the whiskers and the box plots show the interquartile ranges delimited by the 25$^{th}$ percentile (first quartile or Q1) and the 75$^{th}$ percentile (third quartile or Q3). The median is shown by a white dot inside the box plot, and the outliers are outside the whiskers, thus beyond the minimum and maximum values. The outliers were identified using a method that is a function of the interquartile range, as implemented in the Seaborn library in Python. Therein, points which are outside of the 25$^{th}$ and 75$^{th}$ percentile by over 1.5-fold the interquartile range, were labelled as outliers. For the risk analysis, worst-case scenarios can be defined at the maximum resource requirement values, illustrated by the maximum (100th percentile or Q4) top whisker. As shown by the probability distribution, there is a very small chance for this worse-case scenario to materialize based on this analysis. However, even in this worst-case scenario the benefits of establishing new production capacity based on all three platform technologies outweigh the costs by several orders of magnitude when considering the mortality, healthcare burden of the COVID-19 pandemic and economic decline. It is estimated that the pandemic has cost the global economy over 10 trillion USD [31], and the UN projects that the COVID-19 pandemic will reduce the global economy by a further 8.5 trillion USD over a 2-year period [32]. These substantial detrimental impacts of the COVID-19 pandemic can be avoided by comparatively small stimuli in the form of capital and operating costs, ranging between several  hundred million toa few billion USD, as shown in **Figure 5A** and **5B**, respectively. Based on these expenses the total DS production capacity shown in **Figure 5C** can be built to produce the number of batches (**Figure 5D**) required to meet the current shortfall. However, it is worth noting that such investments have to be made ideally in advance, or as soon as possible, considering the years-long timescale required to build such vaccine manufacturing capacity [10]. If vaccine production capacity is built based on platform technologies, such as the RNA platforms and the AVV platform, the resulting facilities could be used for producing a wide range of vaccines over their lifetime.

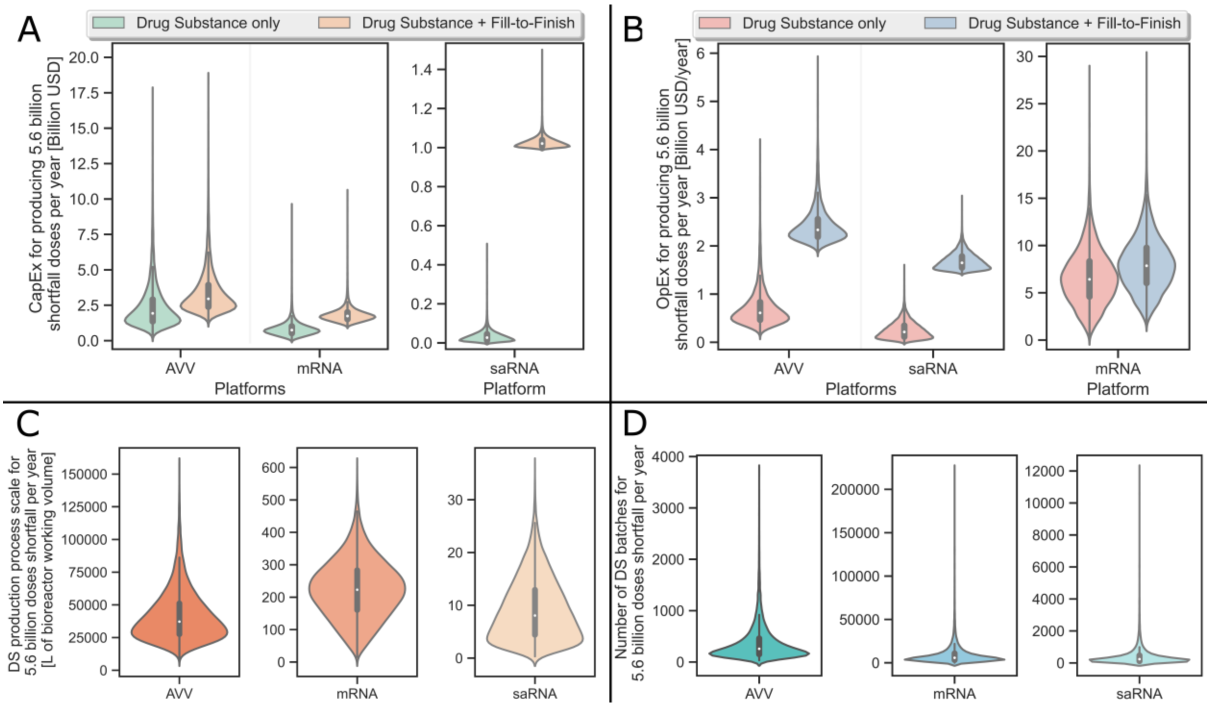


**Figure S5**. Violin plots showing the global sensitivity analysis of the resource requirements for producing the 5.6 billion shortfall COVID-19 vaccine doses within a year using the AVV, mRNA and saRNA production platforms combined with conventional liquid fill-to-finish. The inputs and their ranges used for this global sensitivity analysis are shown in Table 1 in the main text. In the centre of the violin plots, box and whisker plots are shown with the median values indicated by the white dots. **A**. Operating costs (OpEx) required to produce the 5.6 billion shortfall doses per year of AVV, mRNA and saRNA vaccine DS and drug product. It was assumed that AVV vaccine is filled into 10-dose vials, whereas the mRNA and saRNA vaccine is filled into 5-dose vials. **B**. Capital costs (CapEx) required to produce the 5.6 billion shortfall doses per year for the vaccine DS and drug product using the three platform technologies. AVV vaccine fill-to-finish was modelled based on 10-dose vials, whereas the mRNA and saRNA vaccine fill-to-finish was modelled based on 5-dose vials. **C**. Production process scales required to produce the 5.6 billion shortfall doses of DS per year using the AVV, mRNA and saRNA vaccine production platforms. The scale of the production process is represented by the working volume in the bioreactor and the entire process is scaled based on the mass balances proportionally to the bioreactor working volume. **D**. Number of batches required to produce the 5.6 billion shortfall doses of DS per year using the AVV, mRNA and saRNA vaccine production platforms. The box and whisker plots show the minimum and maximum values, except outliers, with the ends of the whiskers; the 25th and 75th percentiles with the top and bottom of the boxes; and the median in the central part of the box.

The resource requirements for producing the annual boost vaccinations are illustrated using the violin shown in **Figure S6**. These results were computed based on global sensitivity analysis. For this, linear extrapolation was assumed which was considered to be a reasonable approximate for scaling out, as these technologies were already modelled at large commercial production scales. These median values are shown in the centre of the box-and-whisker plots. For risk analysis, a worst-case scenario can be assumed based on the maximum resource requirements shown by the ends of the whiskers. The probability of these worst-case scenarios is low, as indicated by the probability distribution represented by the width of the violin plots. Thus, the negative impact of COVID-19, which is estimated at several to tens of trillions of USD [31,32] can be minimized with a relatively small capital investment and operating costs ranging between hundreds of millions of USD to billions of USD. These CapEx and OpEx ranges, shown in **Figure 6A** and **6B**, respectively, will then enable to build facilities with a total DS production capacity shown in **Figure 6C** and run the number of production batches shown in **Figure 6D** necessary to meet annual demand.

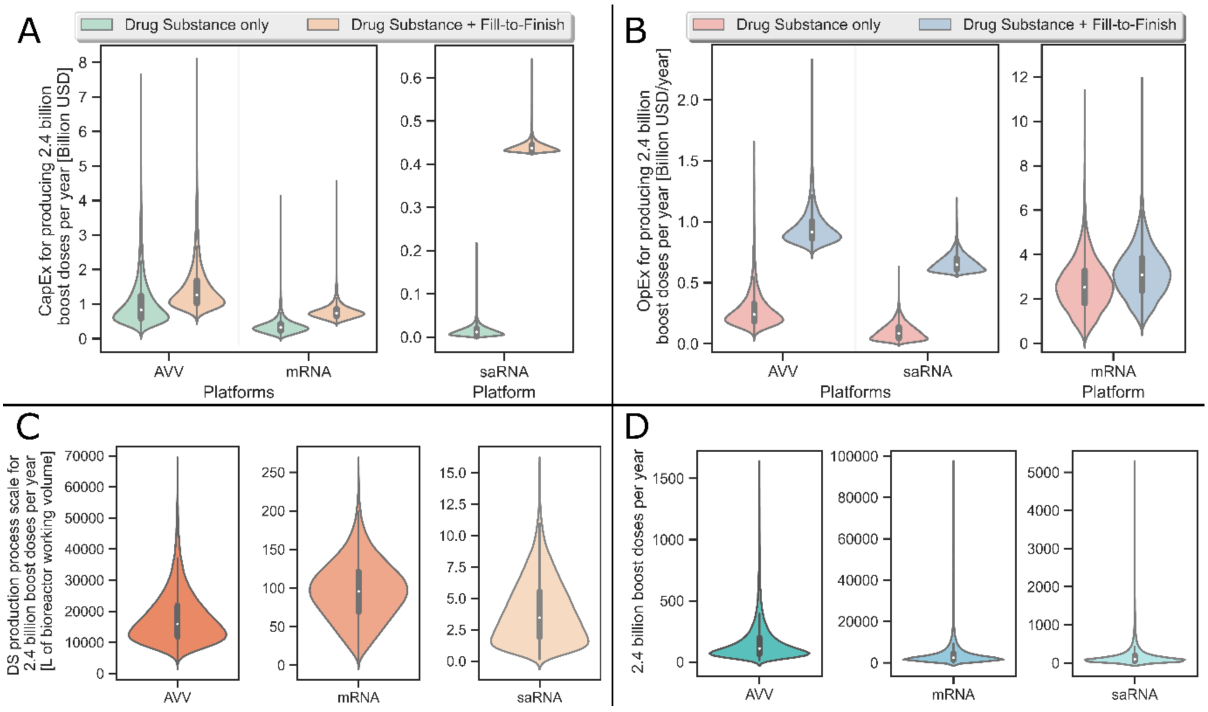


**Figure S6**. Violin plots showing the global sensitivity analysis of the resource requirements for producing the 2.4 billion annual boost COVID-19 vaccine doses per year using the AVV, mRNA and saRNA production platforms combined with conventional liquid fill-to-finish. The inputs and their ranges used for this global sensitivity analysis are shown in Table 1 in the main text. In the centre of the violin plots, box and whisker plots are shown with the median values indicated by the white dots. **A**. Operating costs (OpEx) required to produce the 2.4 billion boost doses per year of AVV, mRNA and saRNA vaccine DS and drug product. It was assumed that AVV vaccine is filled into 10-dose vials, whereas the mRNA and saRNA vaccine is filled into 5-dose vials. **B**. Capital costs (CapEx) required to produce the 2.4 billion boost doses per year for the vaccine DS and drug product using the three platform technologies. AVV vaccine fill-to-finish was modelled based on 10-dose vials, whereas the mRNA and saRNA vaccine fill-to-finish was modelled based on 5-dose vials. **C**. Production process scales required to produce the 2.4 billion boost doses of DS per year using the AVV, mRNA and saRNA vaccine production platforms. The scale of the production process is represented by the working volume in the bioreactor and the entire process is scaled based on the mass balances proportionally to the bioreactor working volume. **D**. Number of batches required to produce the 2.4 billion boost doses of DS per year using the AVV, mRNA and saRNA vaccine production platforms. The box and whisker plots show the minimum and maximum values, except outliers, with the ends of the whiskers; the 25th and 75th percentiles with the top and bottom of the boxes; and the median in the central part of the box.

### 2. Supplementary Methods

#### 2.1. Techno-economic modelling

AVV, mRNA and saRNA DS production (primary manufacturing) modelling as well as drug product manufacturing (fill-to-finish, secondary manufacturing) modelling has been carried out using SuperPro Designer Version 11, Build 2 from Intelligen, Inc. The input parameters and assumptions for DS and drug product techno-economic modelling in SuperPro Designer are listed in **Table S3** below. The time between consecutive batches (cycle slack time) was set to 2 hours in all production process models. All production processes were modelled to operate 335 days per year. The number of campaigns per year was set to 1 in all the DS and drug product manufacturing models. The operating costs (OpEx) include the annualized capital costs (CapEx), calculated using the straight-line depreciation method. The CapEx was included separately from the annualized OpEx to describe the financial resources required to construct facilities. The labour cost for DS production processes (operated in batch mode) was calculated using the detailed labour estimate, in function of the basic labour rate, benefits, operating supplies, supervision cost and administration cost. The labour cost for fill-to-finish processes (operated in continuous mode) was calculated using the lumped labour estimate.

**Table S3.** Input parameters and assumptions used for techno-economic modelling in SuperPro Designer.

| Parameter use | Parameter class | Parameter name | Value | Unit |
|---|---|---|---|---|
| CapEx calculation | Direct Cost (DC) | Piping Cost | 35 | % of TEPC |
| | | Instrumentation Cost | 40 | % of TEPC |
| | | Insulation Cost | 03 | % of TEPC |
| | | Electrical Facilities Cost | 10 | % of TEPC |
| | | Buildings Cost | 250 | % of TEPC |
| | | Yard Improvement Cost | 15 | % of TEPC |
| | | Auxiliary Facilities Cost | 40 | % of TEPC |
| | | Unlisted Equipment Purchase Cost (UEPC) | 30 | % of TEPC |
| | | Unlisted Equipment Installation Cost | 50 | % of UEPC |
| | Indirect Cost (IC) | Engineering Cost | 25 | % of DC |
| | | Construction Cost | 35 | % of DC |
| | Other Cost (OC) | Contractor's Fee | 5 | % of (IC + DC) |
| | | Contingency | 10 | % of (IC + DC) |
| | Miscellaneous | Working Capital – to cover expenses for | 30 AVV;<br>10 mRNA & saRNA<br>30 fill-to-finish | days |
| | | Start-up and Validation Costs | 30 | % of DFC |
| | | Up front R&D | 0 | US$ |
| | | Up front royalties | 0 | US$ |
| OpEx calculation | Facility dependent | Maintenance: equipment specific multipliers | | |
| | | Depreciation: contribution from each equipment's undepreciated purchase cost | | |
| | | Insurance | 1 | % of DFC |
| | | Local taxes | 2 | % of DFC |

| | | | | |
|---|---|---|---|---|
| | | Factory expenses | 5 | % of DFC |
| | Labour | Basic operator labour rate (BOLR) | 20 | USD × hour$^{-1}$ |
| | | Benefits factor | 40 | % of BOLR |
| | | Operating supplies factor | 10 | % of BOLR |
| | | Supervision factor | 20 | % of BOLR |
| | | Administration factor | 60 | % of BOLR |
| | | Lumped operator labour rate | 75 | USD × hour$^{-1}$ |
| | | Adjusted basic operator labour rate* | 46 | USD × hour$^{-1}$ |
| | | Direct labour time utilization - batch | 60 | % |
| | | Direct labour time utilization - continuous | 70 | % |
| | Lab, QC, QA | Laboratory, quality control, quality assurance | 45 | % TLC |
| | Utilities | Standard electricity | 0.1 | US$ × (kW×h)$^{-1}$ |
| | | Chilled water | 0.4 | US$ × tonne$^{-1}$ |
| | | Cooled water | 0.1 | US$ × tonne$^{-1}$ |
| | | Steam | 12 | US$ × tonne$^{-1}$ |
| | Miscellaneous | Fixed R&D | 0 | US$ × year$^{-1}$ |
| | | Variable R&D | 0 | US$ × g MP$^{-1}$ |
| | | On-going process validation | 0 | US$ × year$^{-1}$ |
| | | Other fixed | 0 | US$ × year$^{-1}$ |
| | | Other variable | 0 | US$ × g MP$^{-1}$ |
| Overall economic evaluation | Time valuation | Construction period | 20 | months |
| | | Start-up period | 4 | months |
| | | Project lifetime | 20 | years |
| | | Inflation | 4 | % |
| | | NPV interest - Low | 7 | % |
| | | NPV interest - Medium | 9 | % |
| | | NPV interest - High | 11 | % |
| | Financing | Loan interest for DFC | 9 | % |
| | | Loan interest for working capital | 12 | % |
| | | Loan interest for up front R&D | 12 | % |
| | | Loan interest for up front royalties | 12 | % |
| | | Loan period for DFC | 10 | years |
| | | Loan period for working capital | 6 | years |
| | | Loan period for up front R&D | 6 | years |
| | | Loan period for up front royalties | 6 | years |
| | | DFC outlay for 1$^{st}$ year | 30 | % of DFC |
| | | DFC outlay for 2$^{nd}$ year | 40 | % of DFC |
| | | DFC outlay for 3$^{rd}$ year | 30 | % of DFC |
| | | DFC outlay for 4$^{th}$ year | 0 | % of DFC |
| | | DFC outlay for 5$^{th}$ year | 0 | % of DFC |
| | | Straight line depreciation period | 10 | years |
| | | Salvage value | 5 | % of DFC |
| | Production level | Operating capacity for each year | 100 | % |
| | | Product failure rate | 5 | % |
| | | Disposal cost | 0 | US$ × g MP$^{-1}$ |
| | Miscellaneous | Income tax | 40 | % |

| | | | | |
|---|---|---|---|---|
| | | Fixed advertising and selling expenses | 0 | US$ × $year^{-1}$ |
| | | Variable advertising and selling expenses | 0 | US$ × g $MP^{-1}$ |
| | | Variable running royalty expenses | 0 | US$ × g $MP^{-1}$ |

Abbreviations used in Table S3: CapEx – capital expenditure; OpEx – operating expense; TEPC – total equipment purchase cost; UEPC – unlisted equipment purchase cost; DFC – direct fixed capital; DC – direct cost; IC – indirect cost; OC – other cost; TLC – total labour costs; BOLR – basic operator labour rate; g MP – gram of main product.

*calculated based on benefits, operating supplies, supervision cost and administration cost.

#### 2.2. Sensitivity analysis

Global sensitivity analyses were conducted using SobolGSA Version 3.1.1 available from Imperial College London [33]. SobolGSA generated 10,000 samples within a given set of ranges and distributions of input variables, as indicated in Table 1 in the main text. These 10,000 samples were generated using Sobol sequences, which is one of the quasi-random low-discrepancy sequences [34–37]. These 10,000 quasi-randomly generated combinations of input variables were first sent to MatLab in the form of a matrix. MatLab then called functions in Excel VBA and Excel VBA changed the modelling inputs in the SuperPro Designer model file and started the 10,000 simulations to calculate mass and energy balances and then to perform the economic evaluations. After SuperPro Designer simulated the 10,000 process models, the results were reported to Excel VBA and stored in an Excel file. This Excel file was read by MatLab and the data from the 10,000 simulation results was transferred to SobolGSA. SobolGSA used random-sampling high dimensional model representation (RS-HDMR) [38,39] to generate a metamodel based on the 10,000 simulation results. Finally, SobolGSA calculated the Sobol indices using the metamodel [40]. In addition, SobolGSA conducted 1,250 simulations to test the results predicted by the metamodel. In order to establish the link between SobolGSA and SuperPro Designer, the Component Object Model (COM) interface was employed. The COM interface exchanged data through the path of SobolGSA-MatLab-Excel VBA-SuperPro Designer. For this, MatLab R2020a and Excel from MS Office 365 Enterprise was used.

The input variables used for the variance-based global sensitivity analysis are shown below in **Table S4**. The input value ranges and consequently the uncertainty is slightly lower in case of the AVV platform compared to the RNA platform as the AVV platform is a more established technology. Variations and uncertainties in the downstream purification losses were modelled by varying the titres/yields in the bioreactor, as variations in titre can serve a proxy for variations/uncertainties in downstream losses in terms of amounts of products produced.

**Table S4**. Inputs variables for sensitivity analysis for mRNA, saRNA and AVV vaccine DS production modelling.

| Mfg. Proc. | I/O | Parameter | Range | Probable Value | Unit | Influencing and determining factors | Scenarios, meaning of the parameter range | Ref. |
|---|---|---|---|---|---|---|---|---|
| mRNA and saRNA vaccine | | Process scale | 0.5 – 50 | 30 mRNA<br>5 saRNA | L | Demand, scale-up optimization | Scale required to meet annual demand, larger scale for mRNA and smaller scale for saRNA | [41–44] |
| | | Process failure rate | 0 – 10 | 5 | % | Complexity of the process | Simple robust production process vs complex less reliable production process | |

| | | | | | | | | |
|---|---|---|---|---|---|---|---|---|
| | | Production titres | 3 – 7 | 5 | g × L$^{-1}$ | Reaction optimization, process development | With and without process intensification/optimization | [41,42] |
| | | 5’ cap analogue cost | 2500 – 10000 | 3000 | US$ × g$^{-1}$ | Scale and supplier purchase price | Supplier selling price and supplier production scale | [45] |
| | | Basic labour rate | 5 – 30 | 20 | USD × hour$^{-1}$ | Location of manufacturing | Manufacturing in low-income vs high-income countries | |
| | | RNA amount per dose | 0.1 – 10 for saRNA<br>5 – 150 for mRNA | 1 for saRNA<br>100 for mRNA | µg × dose$^{-1}$ | Clinical trials | Effective DS amount per dose | [46–55] |
| | | Cost of Lab/QC/QA | 15 – 60 | 40 | % of total labour costs | Costs of analytical methods | Low cost analytical methods (e.g. also using soft sensors) vs high cost analytical methods | |
| AVV vector vaccine drug substance production | Inputs | Process scale | 1000 – 20000 | 2000 | L | Demand, scale-up optimization | Scale required to meet annual demand | |
| | | Process failure rate | 0 – 10 | 5 | % | Complexity of the process | Simple robust production process vs complex less reliable production process | |
| | | Production titres, | 1 – 7 × 10$^{14}$ | 2.5 × 10$^{14}$ | viruses × L of culture$^{-1}$ | Reaction optimization, process development | With and without process intensification/optimization | |
| | | Basic labour rate | 5 – 30 | 20 | USD × hour$^{-1}$ | Location of manufacturing | Manufacturing in low-income vs high-income countries | |
| | | AVV amount per dose | 2.2 × 10$^{10}$ – 6.5 × 10$^{10}$ | 5 × 10$^{10}$ | viruses × dose$^{-1}$ | Clinical trials | Effective DS amount per dose | [56–62] |
| | | Cost of Lab/QC/QA | 20 – 60 | 40 | % of total labour costs | Costs of analytical methods | Low cost analytical methods (e.g. also using soft sensors) vs high cost analytical methods | |

#### 2.3. Data processing and plotting

The data obtained from GSA was processed in Microsoft Excel 365 Enterprise and by using the Pandas (version 1.2.4) data analysis library in Python 3.8.3, alongside basic functionalities of the Python 3.8.3 programming language. The line plots, scatter plots and bar/column charts were generated using Microsoft Excel 365 Enterprise. The violin plots were generated using the Seaborn (version 0.11.1) data visualization library in combination with Matplotlib (version 3.3.4) plotting library, both in Python 3.8.3. The violin plots were scaled by “count”, meaning that the relative size of the individual violin plots was determined by the number of data points. The number of data points was constant for all values at 11250 data points (consisting of 10000 GSA simulation results and 1250 SobolGSA test results, as described above in section “2.2. Sensitivity analysis”). The Kernel density estimation (KDE) setting for generating violin plots were left at the default values, with the bandwidth set to “Scott” (based on Scott's rule of thumb [63]). The cut (distance, in units of bandwidth size) was left at the default value of 2 for all violin plots in the SI document. The cut was set to 0 for all violin plots in the main body of the manuscript to limit the longitudinal spread of the violin plots to the data.

Inside the violin plots, box-and-whisker plots are also shown. Therein, similarly to conventional box-and-whisker plots, the minimum (0th percentile or Q0) and maximum (100th percentile or Q4) values are represented using the extremities of the whiskers. The 25th percentile (first quartile or Q1) and the 75th percentile (third quartile or Q3) values are shown by the two ends of the boxes. Thus, the length of the boxes represents the values between the 75th percentile (third quartile or Q3) and 25th percentile (first quartile or Q1), this range/difference is also known as the interquartile range. The median is shown by a white dot inside the box plot. The outliers are outside the whiskers, thus beyond the minimum and maximum values. The outliers were determined using a method that is a function of the interquartile range, as implemented in the Seaborn (version 0.11.1) library. Therein, points which are outside of the 25th and 75th percentile by over 1.5-fold the interquartile range, were labelled as outliers. The width of violin plots represents the probability distributions. The median and the mode of the distributions are often different due to the skewedness of the distributions.

The skewedness of the distribution was so pronounced in some violin plots that the region of interest (region of high probability) was squashed/compressed to accommodate the tail of the skewed distribution in the plotting area. To better visualize the high probability of interest, the top 5% and bottom 5% of the values were removed. Thus, the violin plots were re-built showing only values with magnitudes between 5% and 95%. The top 5% and bottom 5% of the values were removed in the violin plots shown in the main body of the text where the “cut” was also set to zero, as described above. The violin plots shown in the SI document contain 100% of the values (without removing the top 5% and bottom 5%), and the “cut” was also left at the default value of two.

from: http://www.sciencedirect.com/science/article/pii/S0264410X19305328

19. Bosch. Bosch introduces new vial filling and closing machine [Internet]. Manufacturing

Chemist. 2013 [cited 2020 Jul 15]. Available from: https://www.manufacturingchemist.com/news/article_page/Bosch_introduces_new_vial_filling_and_closing_machine/86960

20. Bosch Packaging Technology. Bosch Introduces Vial Filler - MLF 5088 offers an output of 400 vials per minute with IPC [Internet]. Contract Pharma Magazine. 2013 [cited 2020 Jul 12]. Available from: https://www.contractpharma.com/contents/view_breaking-news/2013-03-20/bosch-introduces-vial-filler/

21. Kram T. Email and telephone correspondence with Tim Kram from Rommelag USA, Inc. - June 2020. Evergreen, CO, USA; 2020.

22. Sedita J, Perrella S, Morio M, Berbari M, Hsu J-S, Saxon E, Jarrahian C, Rein-Weston A, Zehrung D. Cost of goods sold and total cost of delivery for oral and parenteral vaccine packaging formats. Vaccine [Internet]. 2018/02/12. Elsevier Science; 2018 Mar 14;36(12):1700–9. Available from: https://pubmed.ncbi.nlm.nih.gov/29449099

23. Jenness E, Walker S. Advantages of single-use technology for vaccine fill-finish operations. PDA J Pharm Sci Technol [Internet]. United States; 2014;68(4):381–3. Available from: https://journal.pda.org/content/68/4/381

24. Rommelag AG. Blow-Fill-Seal Solutions [Internet]. Waiblingen, Germany: Rommelag Kunststoff-Maschinen Vertriebsgesellschaft mbH; 2017. Available from: https://www.rommelag.com/fileadmin/user_upload/Files/CMO/Downloads/EN/Rommelag-Engineering-Products-Brochure-CMO-EN.pdf

25. Rommelag AG. Our bottelpack systems for filling bottles and ampoules [Internet]. Rommelag Kunststoff-Maschinen Vertriebsgesellschaft mbH. 2020 [cited 2020 Oct 28]. Available from: https://www.rommelag.com/en/engineering/products/bfs-aseptic-filling-machines/

26. MEDInstill. Email and teleconference correspondence with experts from MEDInstill - June 2020. New Milford, CT, USA: MEDInstill; 2020.

32. United Nations, Department of Economic and Social Affairs. COVID-19 to slash global economic output by $8.5 trillion over next two years [Internet]. UN. 2020 [cited 2020 Mar

30]. Available from: https://www.un.org/en/desa/covid-19-slash-global-economic-output-85-trillion-over-next-two-years

33. Kucherenko S, Zaccheus O. SobolGSA Software [Internet]. Imperial College London. 2020 [cited 2020 Nov 6]. Available from: https://www.imperial.ac.uk/process-systems-engineering/research/free-software/sobolgsa-software/

34. Sobol' IM. On sensitivity estimation for nonlinear mathematical models. Mat Model [Internet]. 1990;2(1):112–118. Available from: http://www.mathnet.ru/php/archive.phtml?wshow=paper&jrnid=mm&paperid=2320&option_lang=eng

35. Sobol' IM. Sensitivity Estimates for Nonlinear Mathematical Models. Mathematical Modeling and Computational experiment. 1993.

45. TriLink. Telephone conversation with representatives from TriLink, Inc. on 10 April 2020. San

Diego, CA, USA: TriLink; 2020.
